# Quantifying SARS-CoV-2 Omicron variant spread and the impact of non-pharmaceutical interventions in Newfoundland and Labrador, Canada

**DOI:** 10.1101/2025.09.26.25336738

**Authors:** Francis Anokye, Michael WZ Li, Steve Walker, Amy Hurford

**Author notes:** **Author Contributions** **Francis Anokye:** Conceptualization, Methodology, Software, Formal analysis, Visualization, Writing – original draft, Writing – review & editing. **Michael W.Z. Li:** Conceptualization, Methodology, Software, Writing – review & editing. **Steve Walker:** Software, Methodology, Review & editing. **Amy Hurford:** Conceptualization, Methodology, Funding acquisition, Supervision, Writing – review & editing.

## Abstract

The highly transmissible Omicron variant of SARS-CoV-2 caused many infections in Newfoundland and Labrador, Canada, and the fraction of infections that were unreported varied as PCR testing capacity was exceeded and eligibility rules changed. Due to these inconsistencies in the testing rate, we developed a mechanistic model that was calibrated to serological data (Dec 2021–May 2022) to estimate underreporting and understand the impact of non-pharmaceutical interventions on transmission. Our model considers the epidemiology of SARS-CoV-2 spread, natural and vaccine-derived immunity, and the booster dose vaccination campaign that was ongoing in Newfoundland and Labrador. We found that during the early spread of the Omicron variant, when the eligibility for tests that were reported in the provincial counts was less restrictive, three or fewer infections were unreported per reported case. After March 17, 2022, when test eligibility was more restrictive, the underreporting rate increased steadily, with an average of 24.2 infections unreported infections per reported case. We found that Omicron transmission was lower when schools were closed (mean control reproduction number, ℛ_*c*_ = 1.98, 95% CI: 1.58–2.37), higher when open (mean ℛ_*c*_ = 2.71, 95% CI: 2.31–3.11), and of the alert levels, the strictest alert level reduced transmission the most (mean ℛ_*c*_ = 2.23, 95% CI: 1.98–2.53). When underreporting rates vary, the impact of non-pharmaceutical interventions, such as alert level systems and school closures, cannot be determined from reported cases. Our findings highlight the value of combining serological data with modelling to determine the impact of non-pharmaceutical interventions during pandemics when surveillance systems are constrained.

**Author summary:** Reported COVID-19 cases often underestimate the number of infections, especially when testing capacity is exceeded or eligibility rules change. This occurred in Newfoundland and Labrador, Canada, during the spread of the Omicron variant, when testing rates were uneven and many infections were unreported. Our analysis shows that underreporting increased substantially when eligibility for tests that could be reported in the official counts was restricted to high-risk individuals and people who work with high-risk individuals. Given this eligibility change, the underreporting ratio increased from three or less to an average of 24.2 unreported infections per reported case. We used an epidemiological model to account for the many factors, such as asymptomatic infections and vaccination status, that are known to affect infection spread. We found that transmission was lower when schools were closed, higher when schools were open, and of the alert levels, the strictest alert level reduced spread the most. These results highlight that reported cases alone can produce inaccurate results when testing systems are constrained and the rate of testing is variable. Using our approach of combining serological data with modelling enabled us to evaluate the impact of public health measures. These results support knowledge mobilization to explain to the public why particular public health measures are being implemented during an emergency.

## Introduction

COVID-19, caused by the SARS-CoV-2 virus, initiated a public health emergency. As the virus spread, new variants emerged, some of which were variants of concern due to their increased transmissibility and virulence, and the diminished impact of public health measures and vaccines against their spread (1). In November 2021, the Omicron (B.1.1.529) variant (2) emerged and challenged the effectiveness of surveillance programs (3) and non-pharmaceutical interventions (NPIs) (4; 5; 6) due to its high transmission and immune evasion. The effectiveness of surveillance programs and NPIs was challenged even in regions such as the province of Newfoundland and Labrador (NL), Canada that had effectively controlled previous variants and vaccinated a high percentage of their populations (7; 8).

Throughout the pandemic, Canadian provinces and territories required that individuals meeting specific requirements, usually at least one COVID-19 symptom, schedule a Polymerase Chain Reaction (PCR) test to identify SARS-CoV-2 infection. The results of these PCR tests were the basis of reported cases in Canada. During the spread of the Omicron variant, PCR test capacity became insufficient to meet demand, and the provinces and territories adapted by changing the requirements for testing and relying on rapid antigen tests (9), the results of which were usually not reported in official counts (10). These changes increased the proportion of SARS-CoV-2 infections that were not included in the official counts.

Reported cases can produce inaccurate epidemiological curves when the proportion of cases that are unreported changes over time. Serosurveillance can address this challenge by estimating natural infection history more broadly than PCR testing. PCR tests return positive results when viral load is sufficiently high to be detected by the assay (11), while serosurveillance can occur by testing blood samples to identify the antibodies to SARS-CoV-2. Seroconversion refers to an antibody concentration that is high enough to be detected and seroreversion refers to antibody concentrations that can no longer be detected due to waning (occurs over a period of approximately 3–4 months (12)). During the COVID-19 pandemic, serosurveillance was often completed as a cross-sectional survey of blood donors (13). Compared to PCR testing, serosurveillance is less likely to miss asymptomatic infections. Limitations of serosurveillance are whether the blood donor populations are representative of the population, particularly as blood donation has its own eligibility criteria (e.g., age, health status, travel history, and behavioural risk factors), that blood donation centers can be inaccessible to residents of remote and rural populations, and that surveys conducted after antibodies have waned will not detect infections. Serosurveillance identifies both asymptomatic and symptomatic infections and can be compared to reported cases to estimate underreporting rates. From March to May 2020, a longitudinal serosurvey of ten sites in the United States identified 6-24 more infections as compared to official counts (14), and serosurveys in British Columbia, Canada found a substantial increase in underreporting after March 2022 (15).

An epidemic curve where the chance of infections being reported is constant over time is necessary for retrospective studies of control measures to limit infection spread. Such measures are pharmaceutical interventions, i.e. vaccination, and NPIs such as programs of test-trace-isolate, social distancing, capacity limits, mask mandates and recommendations, travel measures and suspending some activities, where school closures are often given particular consideration. Studies show that stricter or earlier implementation of NPIs lowered transmission and hospitalizations even in highly vaccinated populations (5; 16). School closures were often linked with decreases in transmission and hospitalizations due to COVID-19, though the strength of the effect varied by country and timing (5; 16).

In this study, we developed a mechanistic compartmental model stratified by vaccination status, which we calibrated to serological data from December 2021 to May 2022. This approach allowed us to quantify how underreporting varied over time as PCR testing capacity was exceeded and eligibility rules became more restrictive in NL. We estimated the time-varying transmission rate and calculated the reproduction number to evaluate the impact of NPIs. Our analysis quantifies the impact of NPIs on the spread of the Omicron variant in a small, highly vaccinated population that had experienced few SARS-CoV-2 infections from prior variants.

## Methods

### Data

Our analysis covers the period from December 15, 2021 to May 22, 2022 and considers: (i) estimates of infection-induced seroincidence from serological surveys, and (ii) daily reported case counts. Seroincidence estimates were used for model calibration to estimate newly recovered infections, which was used to determine the impact of NPIs and combined with reported cases to estimate underreporting ratios. We interpreted all infections reported during this period as the BA.1 Omicron variant (see Sec. S1.1 in S1 Text for justification). Our study period occurs during the Winter in NL.

### Serological data

From April 2020 to March 2024, the Covid-19 Immunity Task Force (CITF) centralized Canadian SARS-CoV-2 immunity data (17). CITF conducted periodic serological surveys to estimate weekly seroprevalence across Canada using three primary sources: (1) blood donors from Canadian Blood Services and Héma-Québec, (2) anonymized discarded or residual blood samples from provincial laboratories, and (3) participants in CITF-funded research cohorts (13).

### Reported case counts

COVID-19 daily reported cases are publicly available on the Public Health Agency of Canada’s (PHAC’s) website (18), but for this study were provided by the data custodian, the Newfoundland and Labrador Centre for Health Information (NLCHI, now part of NL Health Services – Digital Health), under Health Research Ethics Board approval (2021.013). We applied a left-aligned 7-day moving average to the reported cases. On January 9, 2022, the NL government reported 1009 cases, which were tests that had occurred weeks earlier and had been sent out of the province for analysis. This reporting anomaly affects estimates of the underreporting ratio prior to this date. We note this reporting anomaly specifically, as 1009 cases was the highest daily total reported in NL during the pandemic, and the NL Department of Health and Community Services stated during the January 9 press briefing that these reported cases corresponded to infections that had occurred weeks earlier. No other such reporting anomalies were noted by the NL Department of Health and Community Services during the study period.

### NPIs implemented in NL during the spread of Omicron

NL implemented NPIs of varying strictness, which were described by the COVID-19 Alert Level System (ALS) (19; 20) that had been developed at the beginning of the public health emergency (Sec. S1.2 in S1 Text for a complete summary of the ALS system in NL). Of the alert levels that were implemented during our study period, Alert Level 2 (ALS-2) was the least strict alert level, Alert Level 3 (ALS-3) was of intermediate strictness, and Alert Level 4 (ALS-4) was the strictest, consisting of the lowest capacity limits and more suspended activities. After March 14, 2022, all ALS measures were lifted (No-ALS), as the end of the public health emergency declaration occurred at this time. K–12 schools closures were separate from the ALS system. K–12 schools were closed at times during the study period, partly due to the regular winter break (December 23, 2021 to January 3, 2022), and partly due to recommendations from the Department of Health and Community Services that closed schools for an additional 3 weeks (see Sec. S1.3 in S1 Text for details).

Shortly after the establishment of the Omicron variant in NL, PCR testing capacity was exceeded. As capacity continued to be strained, eligibility rules for PCR tests that were reported in the official counts continued to change. Initially, in December 2021, testing was mandatory for anyone with symptoms or identified as a contact (T1), but by March 17, 2022, eligibility was restricted to only symptomatic individuals that were also high-risk or that could spread infection to high-risk individuals (adults over 60, individuals in congregate settings, pregnant women, immunocompromised individuals, and healthcare workers; T5 see Sec. S1.4 of S1 Text). The periods of ALS levels, K–12 school operational status and changes to PCR testing rules are shown in Fig 1.

**Fig 1.**
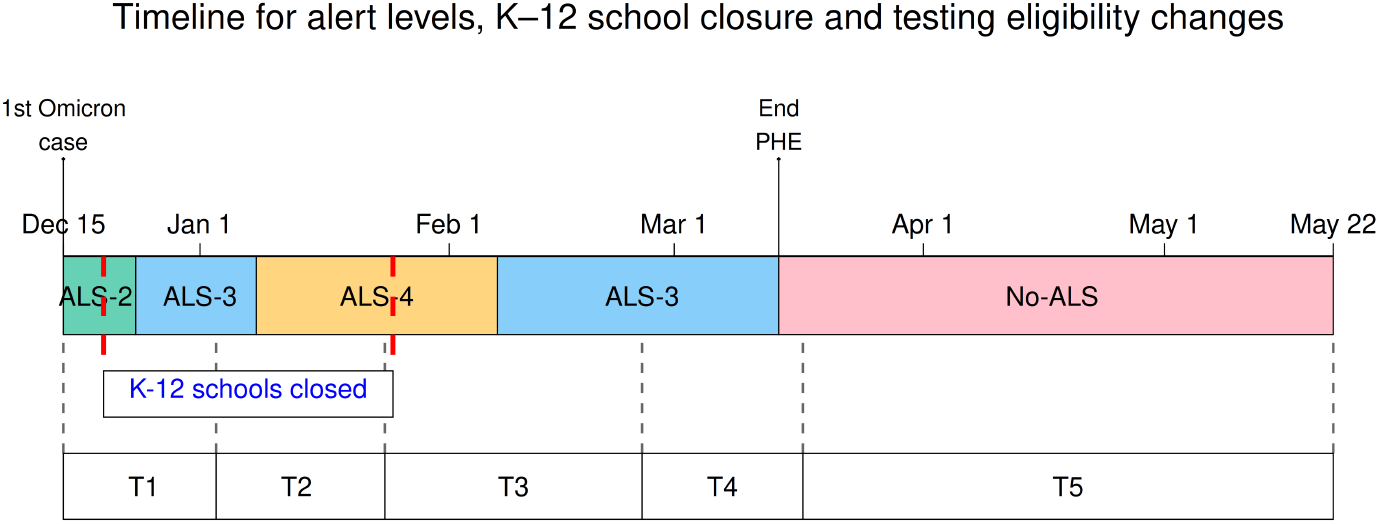
Timeline of alert levels, K–12 school closure, and testing eligibility changes (Dec 15, 2021–May 22, 2022). ‘End PHE’ is the end of the Public Health Emergency declaration. T1–T5 are periods of different PCR test eligibilities. See Sec. 1.2-1.4 in S1 Text) for additional details.

### Vaccination data

Publicly available COVID-19 vaccination data for Newfoundland and Labrador were obtained from the PHAC website (21). These data are reported as cumulative vaccination counts at a weekly resolution.

## Model

To understand the impact of NPIs implemented in NL after the establishment of the Omicron variant, we estimated the time-varying transmission rate using a mechanistic Susceptible-Exposed-Asymptomatic-Infected-Recovered (SEAIR) compartmental model (Fig 2; model equations are described in Sec. S1.5 of S1 Text). Our model is similar to the structures in earlier studies (22; 23; 24) and is stratified into three population cohorts according to vaccination status (we do not distinguish between vaccine types e.g., mRNA vs. live attenuated or brand-specific products), where each cohort has its own SEAIR compartments. The vaccination cohorts are individuals who are: unvaccinated or partially vaccinated (i.e., one dose from a two dose primary series; denoted with a subscript 1); fully vaccinated (i.e., have received the primary series of a Health Canada-approved COVID-19 vaccine, denoted with a subscript 2); booster-vaccinated (i.e., have received additional shots (one or more) to complement the primary series, denoted with a subscript 3).

**Fig 2.**
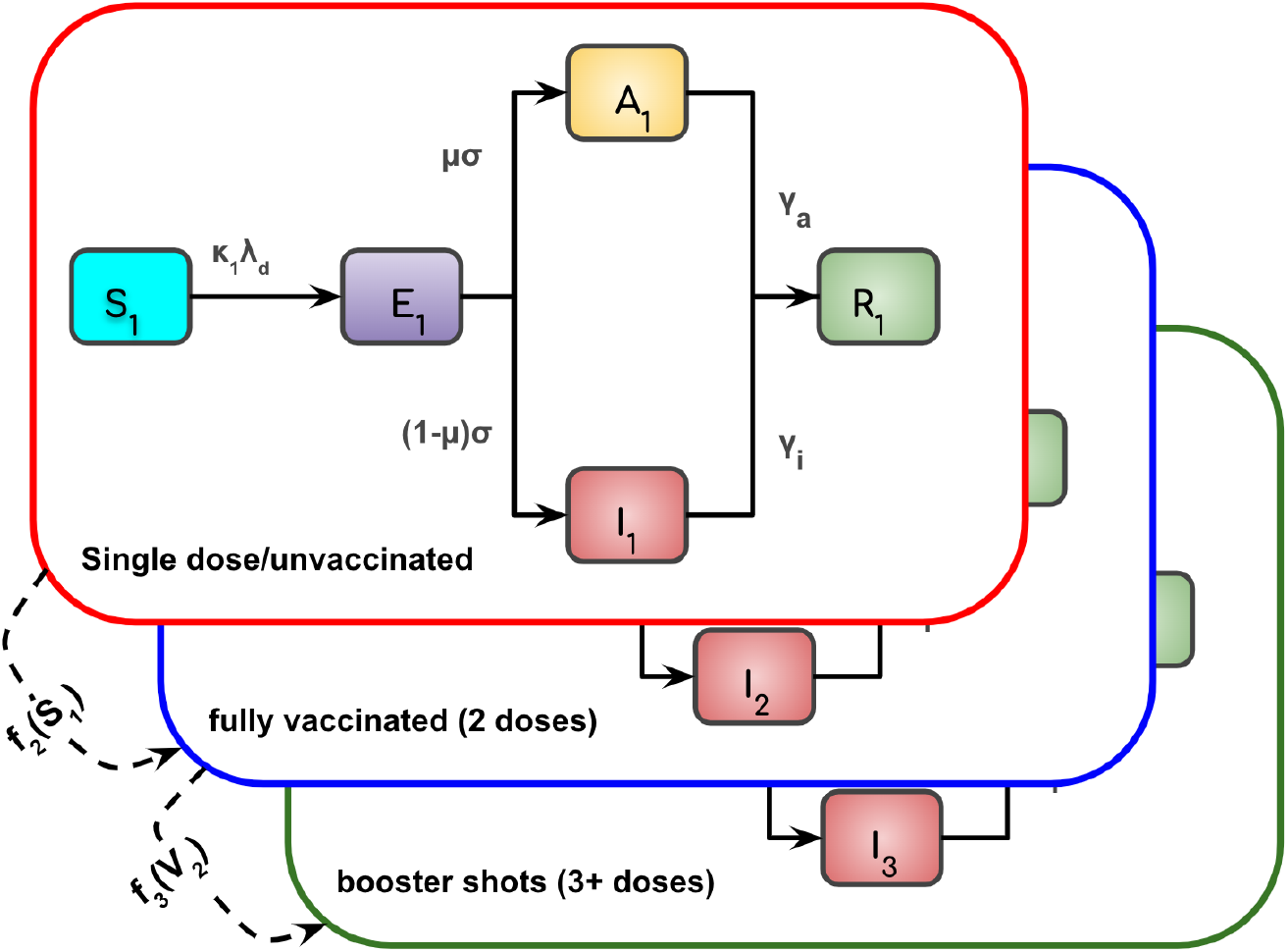
The SEAIR model with three vaccination cohorts. The vaccination cohorts are: unvaccinated/single-dose (cohort 1; red), fully vaccinated (cohort 2; blue), and booster-vaccinated (cohort 3; green). Each cohort contains susceptible, exposed, infectious (symptomatic and asymptomatic), and recovered compartments. Solid arrows show disease progression; dashed arrows show vaccination transitions *f*_2_(*S*_1_) (cohort 1 to cohort 2) and *f*_3_(*V*_2_) (cohort 2 to cohort 3).

We assumed that all cohorts mix equally and have the same contact rates. Asymptomatic infections are scaled by a constant, *ζ*, to reduce the chance of infection given a contact relative to contact with a symptomatic individual. The combined contributions from infectious individuals of all types and from all cohorts are defined by the force of infection, *λ*_*d*_(*t*), mathematically expressed as

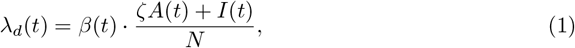

where *β*(*t*) is the estimated time-varying transmission rate defined by Eqn 2 in the next section, *A*(*t*) = *A*_1_(*t*) + *A*_2_(*t*) + *A*_3_(*t*) is the total number of asymptomatic individuals and *I*(*t*) = *I*_1_(*t*) + *I*_2_(*t*) + *I*_3_(*t*) is the total number of symptomatic individuals, where the total is the sum taken across all cohorts. We do not include population demographics such as births and natural deaths due to the short period of the study. Also, we allow only individuals in the susceptible compartments to receive vaccination and transition to another vaccination cohort, because vaccination during infection is rare and unlikely to have any impact at the population level.

Descriptions of model parameters and their values are given in Table 1. Fitted parameters were specified with weakly informative priors (25; 26) centered on defined literature values and complete details are provided in Sec. S1.6 of S1 Text. As there was an active booster dose vaccination campaign during the study period, we modelled individuals moving between the susceptible compartments of the vaccination cohorts at rates that represent the PHAC vaccination data((21); see Sec. S1.7 in S1 Text). We completed a sensitivity analysis to understand the impact of parameter estimate accuracy on our conclusions as described in Sec. S1.8 in S1 Text.

**Table 1.** Model parameters. Fixed parameters are estimated from published sources or data. Fitted parameters with the 95% credible intervals are reported in the ‘Value’ column and assume Gaussian priors with the assumed 95% prior intervals stated in the ‘Details’ column. These fitted parameters are estimated during model calibration (see additional details in Sec. S1.6 in S1 Text).

| Symbol | Definition | Value | Details |
| --- | --- | --- | --- |
| **Fixed parameters** |  |  |  |
| $N$ | Total population | 510,550 | (27) |
| $\mu$ | Proportion symptomatic | 0.676 | (28) |
| $\zeta$ | Relative infectiousness of asymptomatic cases ($0 \leq \zeta \leq 1$) | $\zeta = 0.75$ | (29) |
| $\kappa_1$ | Susceptibility, unvaccinated | $\kappa_1 = 1.0$ | reference value |
| $v_2(t), v_3(t)$ | Daily vaccination rates (2-dose & booster) | vaccination data | Sec. S1.7 in S1 Text |
| **Fitted parameters** |  |  |  |
| $\sigma$ | Rate from exposed to infectious | $0.356 \text{ days}^{-1}$ (CrI: [0.243, 0.469]) | [1/4, 1/2] $\text{days}^{-1}$ (30) |
| $\gamma_a$ | Recovery rate, asymptomatic | $0.106 \text{ days}^{-1}$ (CrI: [0.090, 0.123]) | [1/11, 1/8] $\text{days}^{-1}$ (31) |
| $\gamma_i$ | Recovery rate, symptomatic | $0.144 \text{ days}^{-1}$ (CrI: [0.124, 0.164]) | [1/8, 1/6] $\text{days}^{-1}$ (31) |
| $\kappa_2$ | Relative susceptibility, 2-dose vaccinated ($0 \leq \kappa_2 \leq 1$) | $\kappa_2 = 0.911$ (CrI: [0.862, 0.960]) | [0.85, 0.96] (32) |
| $\kappa_3$ | Relative susceptibility, booster vaccinated ($0 \leq \kappa_3 \leq 1$) | $\kappa_3 = 0.303$ (CrI: [0.229, 0.376]) | [0.20, 0.40] (32) |
| $\beta(t)$ | Time-varying transmission rate (Eqn 2) | $\beta(t)$ is a component of $\mathcal{R}_c$ (see Eqn 3), which is shown in Figure 5. | Discussed in detail in the Methods subsection ‘Time-varying transmission rate’. |

### Time-varying transmission rate

To capture the changes in transmission over time, we modelled the time-varying transmission rate, *β*(*t*), as a smooth exponential spline:

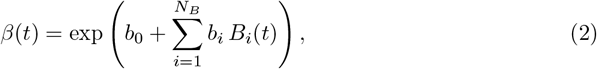

where *t* represents days since *t*_*s*_ − 150 as we begin fitting on *t*_*s*_ = December 15, 2021, and allow for a 150-day burn-in period. We used *N*_*B*_ = 11 natural cubic spline basis functions denoted, *B*_*i*_(*t*), with boundary knots at *t*_*s*_ and *t*_*f*_, where *t*_*f*_ is May 22, 2022. The *b*_*i*_ parameters are spline coefficients that were estimated during the fitting. To regularize the spline and avoid extreme values, we generated the spline coefficients by sampling from the normal distribution with weakly informative priors (25; 26) centered on a biologically plausible value for the log-scale coefficients log(*b*_*i*_), specified as

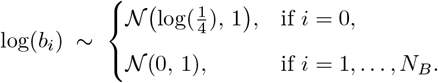

Here, *b*_0_ is the intercept (constant value on the log scale) of the spline basis, and it is estimated once during the model calibration to set the baseline transmission level. The prior, centered on 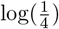, directs the fit toward a realistic baseline transmission rate. The time-varying changes in the transmission were captured by the remaining spline coefficients 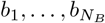.

### Reproduction numbers for levels of the Alert Level System and K–12 school closures

To estimate the transmission rates across a period, [*t*_*a*_, *t*_*b*_], corresponding to an alert level or school closure or opening, we considered all of the daily transmission rate estimates that occurred during this period. We computed the corresponding control reproduction number (see Sec. S1.5 of S1 Text for a derivation) as,

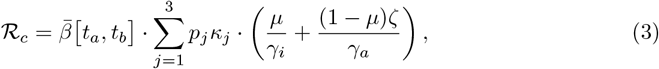

where *p*_*j*_ is the proportion of the population in vaccination cohort *j* and

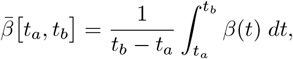

is the mean transmission rate during the period of interest.

The vaccination campaign that was ongoing during our study period affected the reproduction number, so we calculated the reproduction number in a fully susceptible reference population to be able to compare the effect of NPIs without the confounding effect of ongoing vaccination and infection-derived immunity.

If we had not fixed the size of the susceptible population we would have estimated lower reproduction numbers later in the study period, but this would have been partially due to a higher fraction of the population having immunity, which would have prevented us from being able to isolate the effect of different NPIs. When the Omicron variant established in NL, serosurveillance indicated that only ∼2% of the NL population had been infected with SARS-CoV-2, and only a negligible fraction of the NL population had received booster doses, such that the NL population was close to being fully susceptible at the time that the Omicron variant initially established. We reported 95% confidence intervals for all estimated quantities, including the transmission rate and the control reproduction number, ℛ_*c*_, which were calculated using the Wald formula (estimate ± 1.96 × SE, with 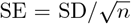, where *n* is the number of observations and SD is the standard deviation).

## Model calibration

We calibrated the compartmental model to the daily infection-induced seroincidence estimates using December 15, 2021 as the start date (denoted *t*_*s*_) and May 22, 2022 as the final date (denoted *t*_*f*_ ). Daily seroincidence was inferred from weekly seroprevalence data using a two-step statistical approach. First, seroprevalence proportions were converted to implied cumulative infections by multiplying by the population size, *N* . A purely statistical B-spline smoothing model was then fitted to these cumulative infections as a function of time using ordinary least squares regression. This spline helps to smooth the seroprevalence data and is distinct from the spline used to parameterize the time-varying transmission rate later used in the SEAIR model. The fitted curve was predicted on a daily scale, and daily seroincidence was obtained as the first difference of the predicted cumulative infections. Model calibration was then performed against these inferred daily infection incidence estimates rather than the cumulative seroprevalence itself. Calibrating to seroprevalence directly may underestimate the uncertainty and give overly narrow confidence intervals (33).

To simulate the epidemic dynamics, we solved the system of ordinary differential equations (see Sec. S1.5 in S1 Text) using a fourth-order Runge–Kutta (34) integration method with daily time steps from *t*_*s*_ − 150 to *t*_*f*_ . We allowed model simulations to begin 150 days before *t*_*s*_ to allow for a gradual buildup of infections such that the proportions of individuals in each compartment at the start of the fitting period, *t*_*s*_, approaches their proportions at equilibrium. At − 150 days before the start of the simulation, *t*_*s*_ − 150, we initialized each vaccination cohort with one individual in each of the exposed, asymptomatic, symptomatic, and recovered states, and set the susceptible populations as 275, 935 (cohort 1) 234, 601 (cohort 2), and 1 (cohort 3). During the 150 day burn-in period, vaccination occurred at rates that represent the PHAC vaccination data, and on *t*_*s*_ = December 15, 2021 this corresponds to vaccination in 82.5% (fully vaccinated, no booster doses) and 3.5% (fully vaccinated with booster doses) of the total NL population (510,543 people), which agrees with the PHAC vaccination data for this date.

We quantified the fit of daily infection-induced seroincidence estimates and model predictions of the newly recovered compartment as a Gaussian likelihood on the log scale beginning from the start of the Omicron wave at *t*_*s*_ = December 15, 2021. Specifically, we let *X*_*j*_ represent the observed seroincidence at time *t*_*j*_ *> t*_*s*_. We obtain the model-predicted number of newly recovered individuals across cohorts at time *t*_*j*_, which is denoted as Δ*R*(*t*_*j*_). For each observation *j*, the likelihood contribution was defined as:

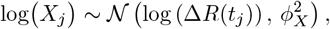

where *ϕ*_*X*_ is an unknown standard deviation parameter estimated from the seroincidence data.

We fitted the model by maximizing the posterior distribution using the macpan2 (35) software package. macpan2 facilitates the estimation of compartmental models in R via Template Model Builder (TMB), a powerful tool for fitting complex latent-variable models. Confidence intervals for parameters, state trajectories, and fitted seroincidence were computed via the delta method as implemented in macpan2.

Due to few community infections occurring initially during Omicron spread in NL, random events in small heterogeneous populations were impacting infection spread, and this could not be reproduced from our model. Therefore, we decided to only report results beginning on January 1, 2022 when infection had spread more substantially and these effects likely no longer persisted. For completeness, we show results beginning from the establishment of the Omicron variant on December 15, 2021, but results from before January 1, 2022 are shaded in grey, and we de-emphasize the interpretation of results from this early period of infection spread.

The code used in this study is archived on Zenodo (36).

## Results

SARS-CoV-2 incidence, which we inferred from model, increased from December 15, 2021 until reaching a peak of 1,854 infections in late April 2022, after which incidence declined (Fig 3). Reported infections peaked twice in early January (677 infections) and in early March 2022 (541 infections). During the testing eligibility periods T1-T4, we estimated that the average underreporting ratio was 1.0, 0.9, 3.0, and 2.7 unreported infections per reported infection, respectively, and the underreporting ratios were relatively constant during this period (Fig 4B). During the testing eligibility period T5, the average underreporting ratio was 24.2 infections unreported for every 1 reported infection and the underreporting ratio increased from the beginning of the implementation of T5, reached a peak of more than 50 unreported infections per reported infection, and then declined to near 40 unreported infections per reported infection at the end of the study period. The T5 period corresponds to when the eligibility for testing was restricted to high-risk symptomatic individuals or people who work with those at high-risk (Sec. S1.4 in S1 Text).

**Fig 3.**
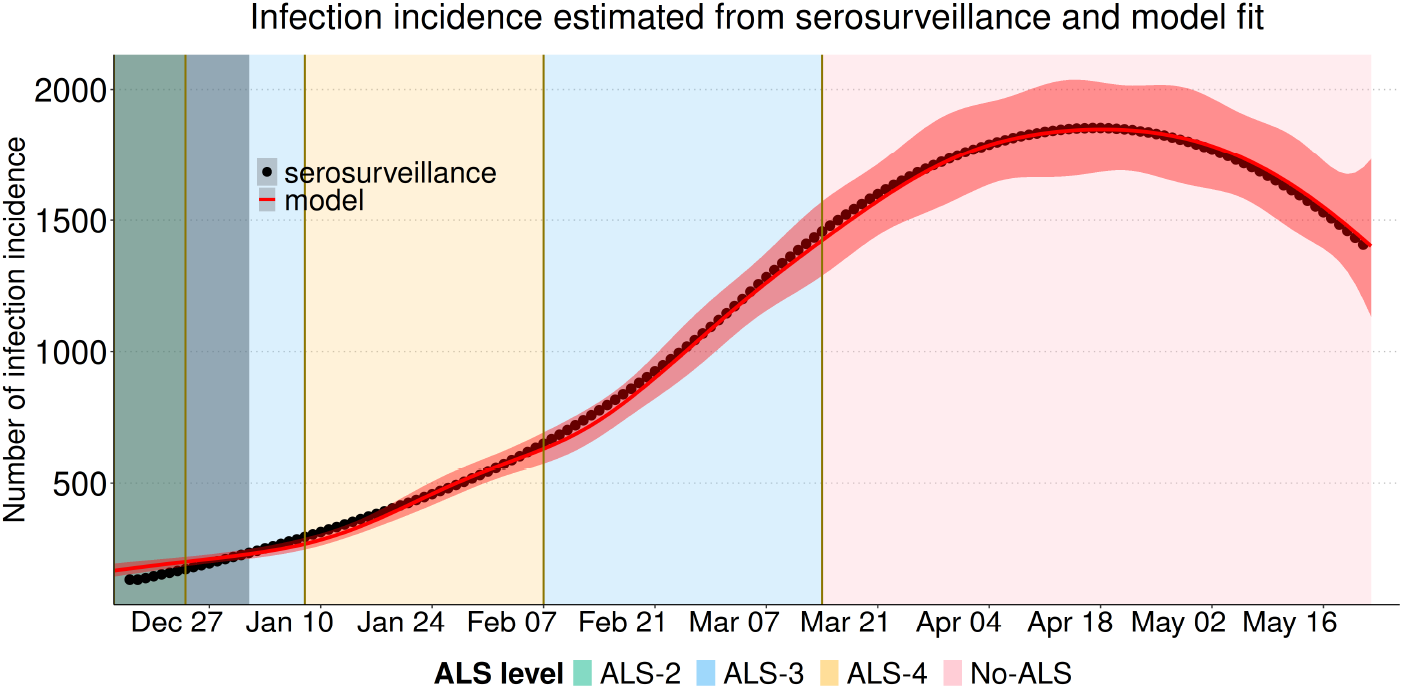
Model fit to infection incidence as calculated from serological prevalence data for December 15, 2021–May 22, 2022. Model-estimated incidence (red line, shaded band is the 95% confidence interval) fit to the seroincidence data (black dots). Background shaded colors indicate different levels of NPIs under the ALS system (from least to most strict): No-ALS (pink); ALS-2 (pale green), ALS-3 (pale blue); and ALS-4 (pale yellow). The solid vertical line indicates the date on which the public health emergency declaration that instituted the ALS was repealed.

**Fig 4.**
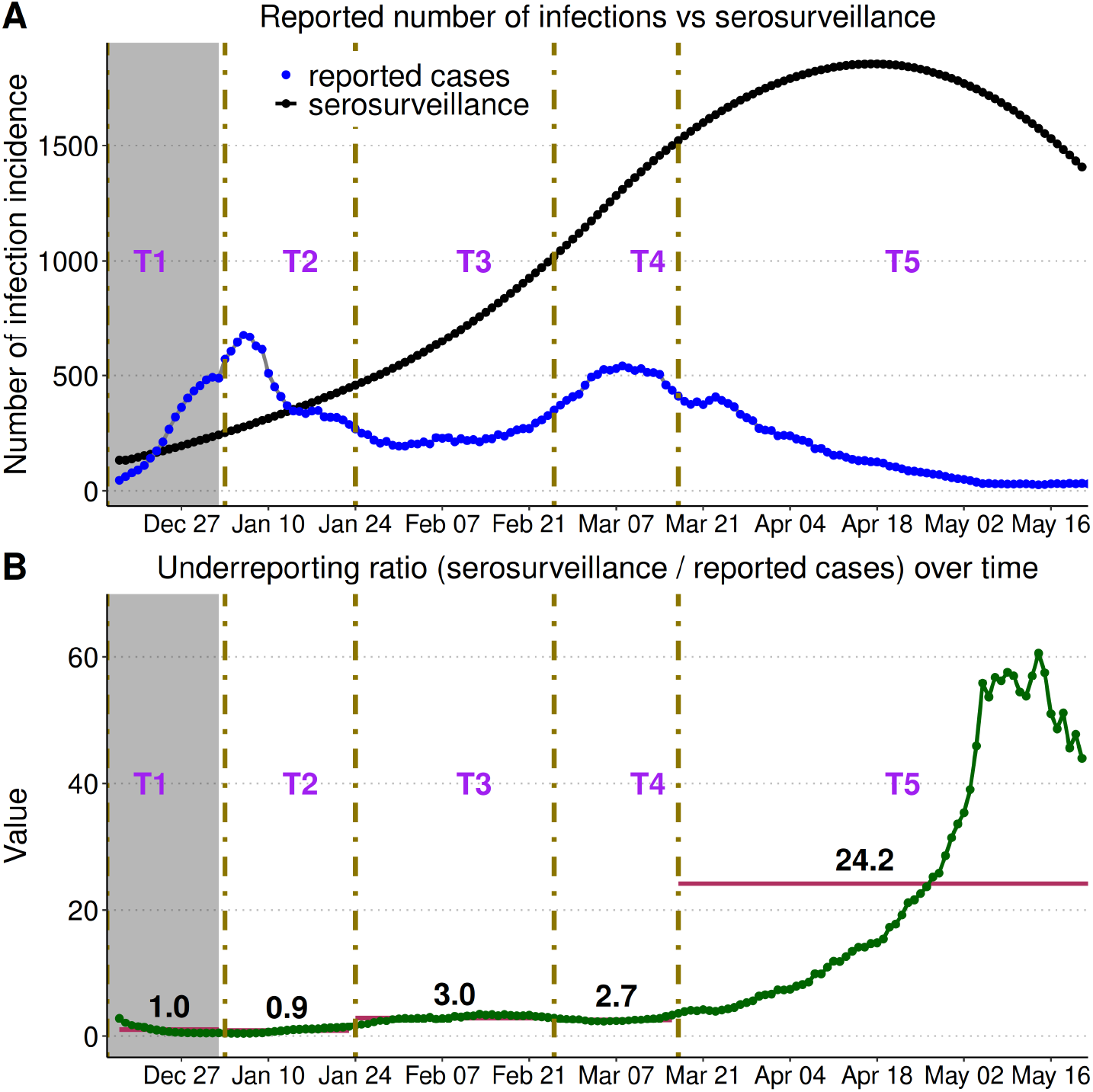
Daily reported cases, incidence inferred from serosurveillence, and the underreporting ratio during the Omicron wave in NL. (A) Incidence as inferred from serosurveillence (black) and reported incidence (blue) with changes in testing eligibility periods (T1–T5) shown as vertical lines. (B) The daily underreporting ratio, calculated as daily incidence estimated from seroincidence divided by daily reported cases, with bars representing the mean ratio associated with each testing period. The grey shaded region is considered unrealiable due this being early in the outbreak.

We used our estimate of the time-varying transmission rate *β*(*t*) to calculate a time-varying control reproduction number (ℛ_*c*_). The control reproduction number increased from around 1.6 at the end of the early period (January 1), leveling off and remaining relatively constant at around 2.5 from late February until the end of the study period (Fig 5). The closure of K-12 schools (Fig 5; vertical red dotted lines) occurs early in the study period and the control reproduction number is lower at this time. The control reproduction number stops increasing when the ALS-4 (pale yellow; Fig 5) is implemented in January. The control reproduction number increases in February when the less strict ALS-3 (pale blue shading; Fig 5) is implemented, and is relatively constant then after at around 2.5, which includes the period when the public health emergency was repealed and the ALS system no longer applied (No-ALS, pink shading; Fig 5). We estimate that at no time was the control reproduction number less than 1, however, it should be noted that we calculate the control reproduction for an unchanging population of all susceptible individuals (see Eqn 3) to enable our comparison of the NPIs. The effective reproduction number would be less than 1 by the end of the study period as indicated by the decrease in incidence shown in Fig 3 due to the immunity that has accumulated in the population.

**Fig 5.**
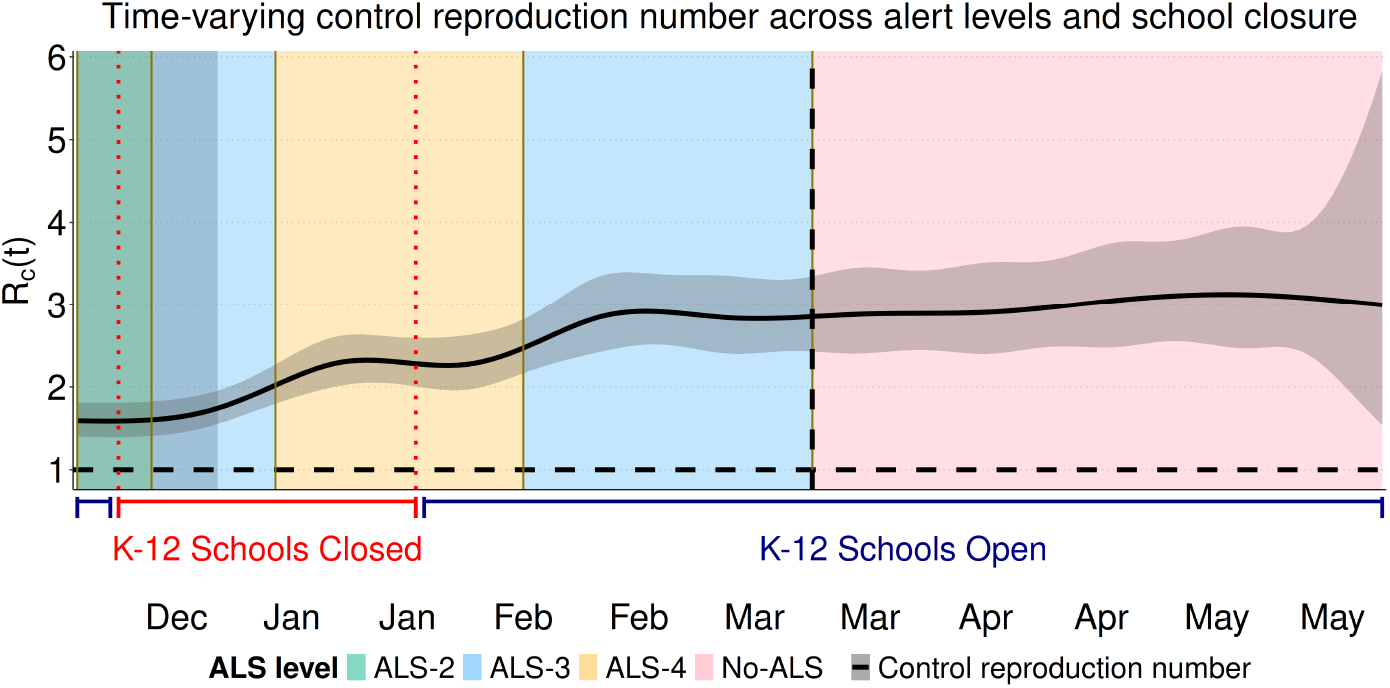
Time-varying control reproduction number during the Omicron wave in Newfoundland and Labrador. (A) The solid black line shows the estimated control reproduction number ℛ_*c*_ and its 95% confidence interval. K–12 schools were closed during the period between the red dotted lines. Background shaded colors indicate different levels of NPIs under the ALS system (from least to most strict): No-ALS (pink); ALS-2 (pale green), ALS-3 (pale blue); and ALS-4 (pale yellow). Grey shading indicate the early period. The black dashed vertical line indicates the date (March 14, 2022) on which the public health emergency declaration was lifted and corresponds to the start of the No-ALS period (pink).

Omicron transmission was found to be related to periods of school closures and the alert levels that were implemented. Periods when schools were closed consistently had lower levels of infection spread (mean ℛ_*c*_ = 1.98, 95% CI: 1.58–2.37), compared to school-open periods (mean ℛ_*c*_ = 2.71, 95% CI: 2.31–3.11; Fig 6A). When ALS evaluation was performed independently of school operational status, the mean control reproduction number increased with decreasing strictness of the alert levels. The lowest control reproduction number values were observed during ALS–4 (2.23, 95% CI: 1.98–2.53; Fig. 6B; pale yellow), higher values were observed during ALS–3 (2.74, 95% CI: 2.18–3.32; Fig. 6B; pale blue bar), and the highest values were observed during the No-ALS phase (3.00, 95% CI: 2.82–3.18; Fig. 6B; pink bar). These estimates indicate that the intensity of transmission increased progressively with the relaxation of the alert levels. We found the most variability in ℛ_*c*_ during ALS–3 (Fig. 5; pale blue), which occurred at two different times, and was implemented for the most number of days.

**Fig 6.**
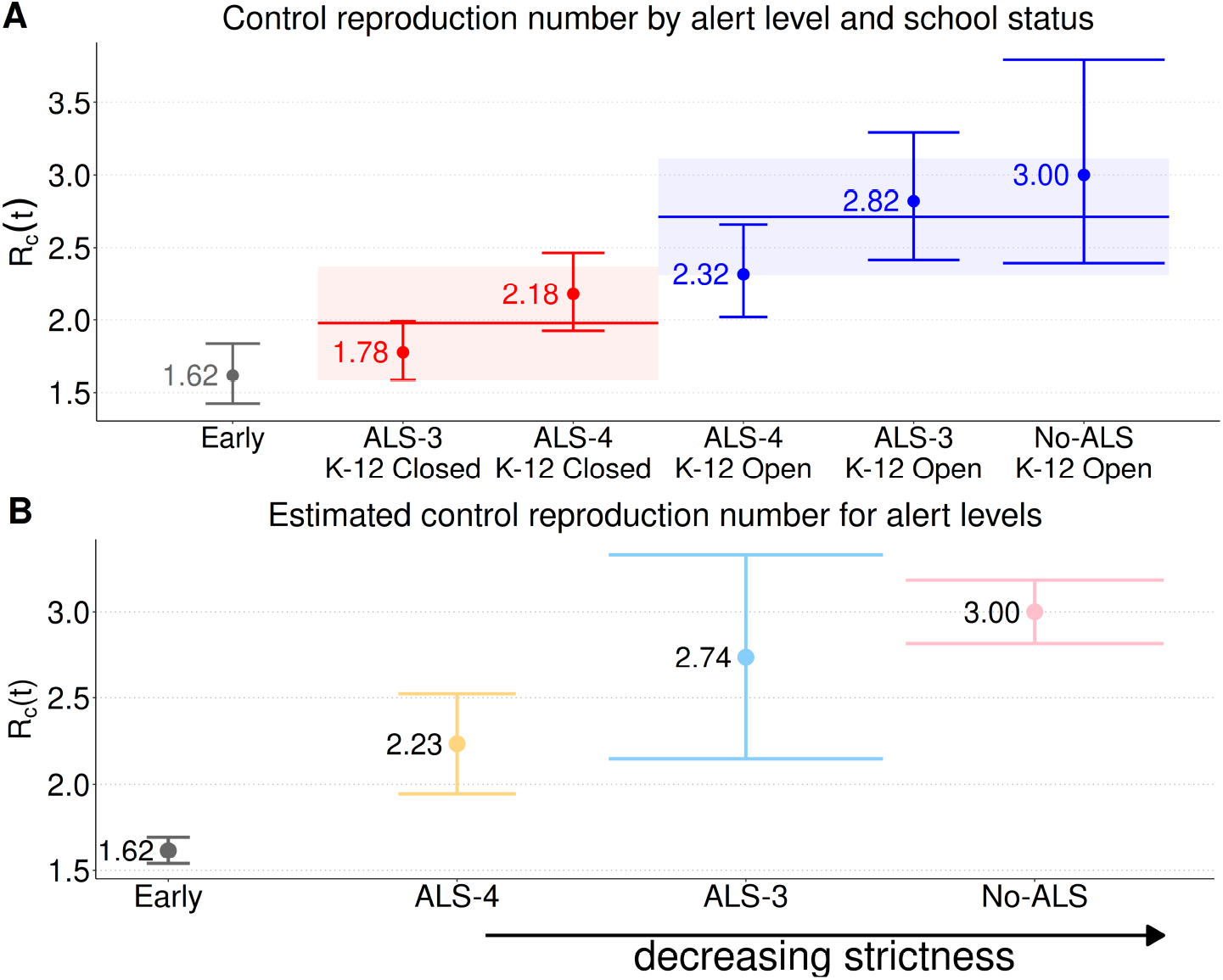
Estimated control reproduction number, ℛ_*c*_, for different school closure and alert level combinations. Points indicate mean estimates, and vertical lines represent 95% confidence intervals. The horizontal width of each confidence interval indicates how long that alert level lasted, with wider bars indicating that alert level covered more days. (A) Alert level and K–12 school status combinations, where the horizontal lines mark the overall mean ℛ_*c*_ for each group: 1.98 [1.58–2.37] for school closures (red line) and 2.71 [2.31–3.11] for schools open (blue line). The control reproduction number, ℛ_*c*_, was consistently lower during K–12 closures and significantly higher after opening. (B) The control reproduction number, ℛ_*c*_, for each alert level irrespective of school operational status. The arrow indicates the direction of increasing strictness of the alert levels.

We performed a sensitivity analysis that considered a range of *µ* and *ζ* values (Sec. S1.8 in S1 Text). We found that the time-varying control reproduction number (ℛ_*c*_) is nearly identical for different values of *µ* and *ζ* given our modelling approach. This is because changes to the fixed values of *µ* and *ζ* are compensated for during the refitting of *σ, γ*_*a*_, *γ*_*i*_, *κ*_2_, *κ*_3_ and *β*(*t*) to the seroincidence data. The overall impact of changing the fixed parameters (*µ* and *ζ*) and refitting the other model parameters is that ℛ_*c*_(*t*), which is a composite measure that combines all of these parameters (Eqn 3), is nearly unchanged (Sec. S1.8 in S1 Text). The values of the fitted parameters are epidemiologically plausible (Table 1) and the posterior distributions of the fitted parameters are similar to the priors (Sec. S1.6 of S1 Text).

## Discussion

Consistent estimation of the number of infections is necessary to evaluate the impact of public health interventions during a pandemic. Our analysis considered serosurveillance data, as in Newfoundland and Labrador (NL), the highly transmissible Omicron variant overwhelmed PCR testing capacity, and testing eligibility changed several times during the study period. During the period when testing eligibility was most restricted, the underreporting ratio increased substantially from three or fewer unreported infections per reported case to an average of 24.2 unreported infections per reported case (Fig. 4B). We also found that K–12 school closure and the strictest alert level (ALS-4) was associated with lower transmission. While NL had successfully controlled the Original, Alpha, and Delta SARS-CoV-2 variants (7; 8), we found that none of the combinations of NPIs implemented during the spread of the Omicron variant resulted in estimates of ℛ_*c*_ < 1 which would correspond to the ability to eliminate the Omicron variant in a wholly susceptible population.

The impact of NPIs implemented in Canada during the COVID-19 pandemic has been estimated by calibrating models to reported cases (37), but during the Omicron wave this was not possible because PCR testing capacity was exceeded in most jurisdictions (9) and the testing rate varied over time, such that the epidemiological curve based on reported cases did not reflect the actual epidemiological dynamics (38). For Ontario, an adjustment to reported cases that applied the testing rate of the most tested group to the entire Ontario population estimated 3.1 unreported cases for every reported case (between March 1, 2020 and September 4, 2022 (38)), and an adjustment based on model calibration to reported cases, hospitalizations, and deaths found 2.2 unreported cases per reported case (January 1–February 9, 2022 (9)). The periods for these estimates are different than for our study, so it is difficult to compare these estimates with our estimates for NL. It is reasonable that we estimate more than 50 unreported infections per reported infection in May 2022 in NL (Fig. 4B), as even higher underreporting ratios are estimated in British Columbia, Canada over a corresponding time period (15).

Potentially explaining our high estimate of the underreporting ratio during T5 is that few Newfoundlanders and Labradorians had been infected with other SARS-CoV-2 variants, and so it is possible that there are a higher number of unreported Omicron infections in NL than for jurisdictions such as Ontario, which had experienced more infections prior to the establishment of the Omicron variant. Across Canada, there was substantial variation in the dates when restrictive PCR eligibility criteria were implemented (Sec. S1.9 in S1 Text) after the establishment of the Omicron variant. NL’s implementation on March 17, 2022 was the latest of all Canadian provinces with the exception of Prince Edward Island, which never limited PCR test eligibility to individuals in high-risk groups and that work with individuals in high-risk groups.

We found that in early January 2022, reported incidence briefly exceeded incidence as inferred from serosurveillance (Fig. 4A), but this has been noted in other studies (39), and has been a characteristic of Canadian serosurveillance data, particularly during periods of, or for provinces with, low infection incidence (see Sec. S1.10 in S1 Text). The alternative method of fitting the model to hospitalization and mortality data could not be applied in NL because hospitalizations and deaths were too few to use for model calibration.

We found that the seroincidence curve for NL has just one peak in mid-April 2022, while reported cases peak in early January and again in early March, with a low occurring in mid-February. These highs and lows in reported cases correspond to periods when particular combinations of alert levels and school closures occurred in NL, such that fitting our model to reported cases would produce substantially different and erroneous conclusions regarding the relative efficacies of the different NPIs. We found that K–12 school closures were associated with lower control reproduction numbers (Fig. 6A) and that ALS-4 had the lowest control reproduction number (Fig. 6B), which is reasonable as these lower reproduction numbers correspond to stricter NPIs.

For comparison of our modelling to other Canadian jurisdictions, we consider the analysis reported in real time for Ontario that would correspond to the period of BA.1 variant spread (40). The serosurveillance data that we used for our study was not available in real time, but hospital ward and Intensive Care Unit occupancy are likely correlated with incidence. In Ontario, the Omicron variant was first detected on November 28, 2021 and analysis showed that hospital ward and Intensive Care Unit occupancy in Ontario each increase to a single peak which occurs in mid-January (40). These graphs of hospital ward and Intensive Care Unit occupancy due to COVID-19 in Ontario have the same shape as our seroincidence graph (Fig. 3), but our seroincidence curve for NL does not peak until 3 months later in mid-April. The later peak in infections in NL suggests that the transmission rate for the BA.1 variant in NL may have been lower than in Ontario.

We estimated the control reproduction number for the Omicron BA.1 variant in NL when no alert levels or school closures were in place at 3.00 (Fig. 6B). An effective reproduction number can be estimated by multiplying our estimate of the control reproduction numbers by an assumed fraction of the population that is susceptible. In Ontario, the effective reproduction number for the BA.1 variant was estimated as 4.01 (41), so if 90% of the NL population is assumed susceptible, our analysis would estimate an effective reproduction number of 2.7 for NL with no NPIs in place. Worldwide, the average basic reproduction number for the BA.1 variant was estimated as 9.5 (42), and because our control reproduction number is calculated for a wholly susceptible population, our estimate of the basic reproduction number in NL is 3.00. There are several reasons why the reproduction numbers for Omicron variant spread in NL would be less than Ontario and/or worldwide averages. Firstly, none of the Original, Alpha, and Delta variants spread very extensively in the NL community, while these variants did spread in Ontario and the countries that are the basis for the reproduction number estimates reported in (42). The Original, Alpha, and Delta variants did not spread in the NL community because public health officials implemented an effective containment strategy (43), but more generally, all the factors that determine why some regions, such as NL, were able to contain the spread of SARS-CoV-2, and others were not, is a subject that requires further study (44).

Shortly after the establishment of the Omicron variant in NL, we observed that the transmission rate was lower than at any other time, irrespective of the NPIs that were implemented (Fig. 5; left-most shaded grey). This may be reasonable, as NL had effectively controlled COVID-19 until the establishment of the Omicron variant, and the public may have exercised greater caution in response to the perceived unprecedented risk due to the establishment of the Omicron variant. Few Newfoundlanders and Labradorians had been infected with SARS-CoV-2 prior to the establishment of Omicron (∼2% as of December 15, 2021), and the high percentage of the NL population that was vaccinated suggests that most of the NL population followed public health directives to reduce the spread of SARS-CoV-2. Shortly after the establishment of the Omicron variant, in the Government of NL public briefings, it was stated that it was no longer possible to contact trace all SARS-CoV-2 infections (43) and that most people will contract the virus at some time (45). This combination of NL’s low infection rate prior to Omicron, compliance with measures to limit the spread of the virus, and the high risk of Omicron variant infection may have resulted in voluntary actions that reduced the infection transmission rate, and this may explain why we estimate low transmission rates, and control reproduction numbers, early in the outbreak.

Among the measures implemented, we observed that K–12 school closures were the most effective in reducing the Omicron variant transmission rate in NL, but these closures also occurred early in the outbreak, and this timing might explain our finding rather than the school closures themselves. Voluntary risk-reduction, uneven adherence to restrictions, and differences in contact patterns between age groups or settings likely influenced transmission, but could not be explicitly captured in our model. The limitation of our study in this aspect could have been mitigated if there were available data describing the NL population’s behavioural response to the establishment of the Omicron variant, which might have occurred through surveys reporting age-structured contacts and attitudes or by measuring contacts and mobility.

Our SEAIR model considered many of the processes known to impact the spread of SARS-CoV-2, but a consistent barrier to our analysis was insufficient data. The reported case data alone are insufficient to determine the impacts of NPIs on Omicron spread due to PCR capacity having been exceeded and changes to PCR test eligibility throughout the study period. We calibrated our model to seroincidence estimates, but these data were not available in real time to support decision-making. Hospitalizations and deaths were too few in NL for these data to be useful for calibration, making other data sources even more important. The vaccination data that we used have reporting gaps in the first weeks of our study, and data describing contacts, mobility, and behaviour were not collected. During the public health emergency, resources were stretched, and this may explain the limited data. Improved data collection and sharing would enable analyses that could more definitely establish why the transmission rate was low after the initial establishment of the Omicron variant and inform public health strategies.

For our study, mathematical modelling was needed to disentangle the effects of the different NPIs implemented in NL, due to their implementation at different times during the Omicron outbreak, because during the study a vaccination campaign was ongoing, and because the periods when different NPIs were implemented were overlapping, such that this combination of factors makes it difficult to estimate the effect of the different NPIs within a simplified analysis framework. Our analysis quantifies the impact of NPIs, which supports decisions (i.e., to enact or discontinue an ALS-4) and provides transparency that builds public trust and improves compliance with public health measures. Improving data collection and supporting modelling analysis of SARS-CoV-2 transmission would benefit future emergency responses.

### S1 Text. Model equations, data and additional results

SEAIR model equations, parameter priors, and descriptions of seroprevalence, reported cases, vaccination uptake, sensitivity analysis, and variant composition data. Includes supplementary figures (BA.1 dominance, reported cases vs. seroprevalence for all Canadian provinces, vaccination uptake) and tables (ALS measures, school closures, PCR testing eligibility, provincial PCR restrictions, and underreporting ratios).

## Data Availability

All data and code used to produce the results presented in the paper and in the
supplementary information are available online at

https://doi.org/10.5281/zenodo.18497732

## Acknowledgments

We thank the Newfoundland and Labrador Centre for Health Information (NLCHI, now part of NL Health Services – Digital Health) for providing access to the data (Health Research Ethics Board reference number 2021.013). We are also grateful to the COVID-19 Immunity Task Force (CITF) for serosurveillance, to the developers of the macpan2 software, and to the public health officials and healthcare workers in Newfoundland and Labrador whose efforts in surveillance and reporting made this analysis possible.

