## Supplementary Information for "Quantifying SARS-CoV-2 Omicron variant spread and the impact of non-pharmaceutical interventions in Newfoundland and Labrador, Canada"

### S1 Text: Supplementary Material

#### S1.1 BA.1 Dominance in Canada

COVID-19 genomic surveillance and variant composition derived from comprehensive whole-genome sequencing (1) indicates that the majority of SARS-CoV-2 infections in Canada during the study period (January 2022 to May 2022) were attributable to the Omicron BA.1 sub-lineages (BA.1, BA.1.1, BA.1.1.10, BA.1.1.16, BA.1.1.18, BA.1.1.6, BA.1.14, BA.1.15, BA.1.17, BA.1.17.2, BA.1.20, BA.1.3). In Fig. S1, we aggregated the BA.1 family as 'BA.1' and all remaining lineages are pooled as 'other lineages'. This suggests that Canadian sequences were dominated by BA.1 and its sub-variants during this period, aligning with global trends observed during the initial Omicron wave. The BA.1 fraction was also reported for Newfoundland and Labrador (2; 3) and the visualization on the CoVaRR-Net website shows that the BA.1 variant takeover occurred more rapidly in NL than in Canada overall. This is reasonable as there were fewer Delta variant cases in NL than other regions in Canada when the BA.1 variant first established. We were not able to access the variant fraction data for NL. Nonetheless, the evidence is compelling that the variant spreading in NL during the period of our analysis was mostly BA.1.

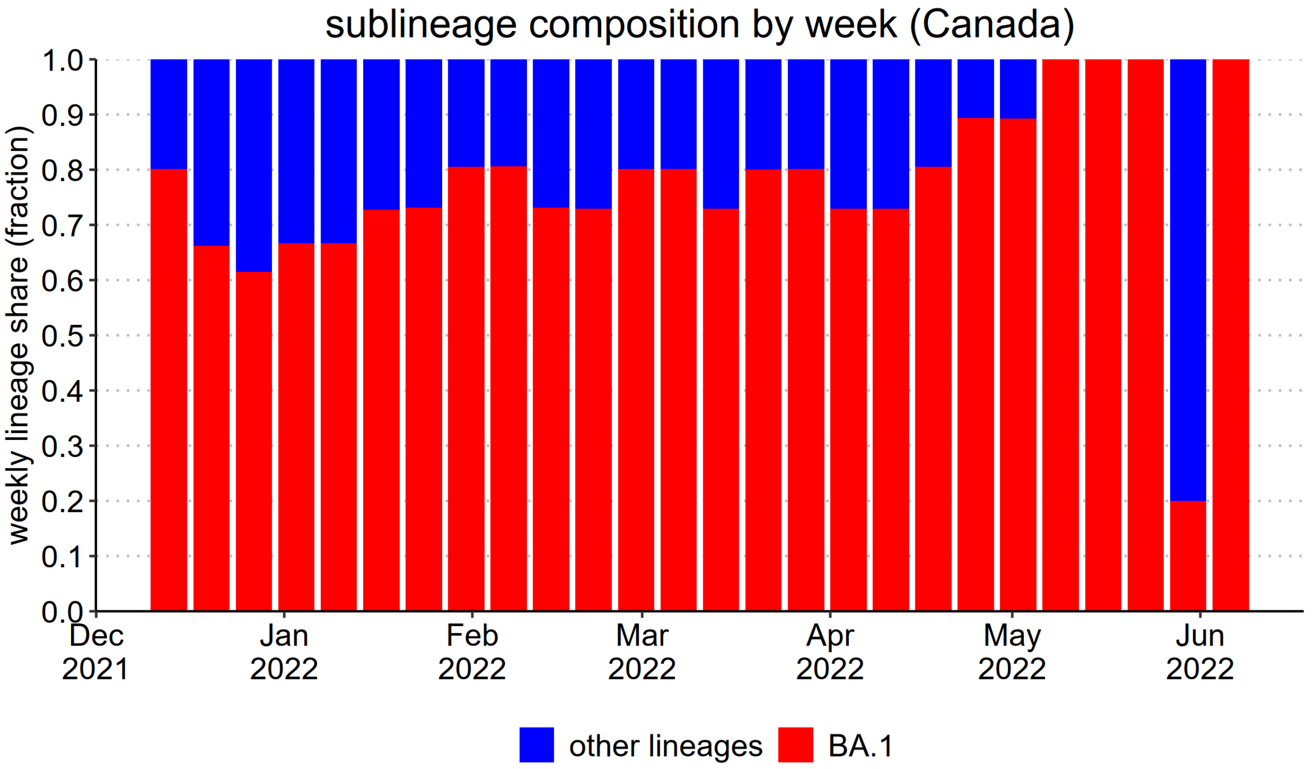


**Fig. S1**: **Omicron BA.1 Sub-lineages in Canada.** Each bar shows the fraction of sequenced cases per week that where the BA.1 variant. Fractions were normalized within week to sum to 1. Data is from (2)

#### S1.2 Alert Level System in Newfoundland and Labrador

The Alert Level System (ALS) is a five-tiered structured framework to gradually lift public health restrictions under the Public Health Emergency in Newfoundland and Labrador (NL) (4; 5). The ALS levels range from the strictest public health measures (Level 5) to the 'Living with COVID-19' (Level 1). Each level represents the degree of restrictions and public health measures necessary to control the transmission of COVID-19 while allowing for the resumption of activities. Alert Level 1 (ALS-1) represented the easing phase, during which withdrawal of long-term public health restrictions was considered. Alert Level 2 (ALS-2) introduced moderate interventions such as indoor capacity limits. Under Alert Level 3 (ALS-3), stricter NPIs were implemented including suspension of some activities and business operations. A relaxed version of Alert Level 3 (ALS-3) phase was reintroduced later, applying targeted restrictions to social and recreational settings while maintaining limited essential services under heightened measures. Alert Level 4 (ALS-4) entailed stricter measures, including closures of high-contact venues and lower gathering limits, with only essential services permitted under enhanced distancing protocols. Alert Level 5 (ALS-5) was not implemented during the spread of the Omicron variant, but was the most restrictive level, involving near-total shutdowns of non-essential services and gatherings to preserve critical operations. No ALS refers to the period that covers the repeal of the public health emergency declaration that instituted the ALS. Under No-ALS, only the public health measures during a non-emergency were implemented. The details of these measures are presented in Table S1.

**Table S1**: COVID-19 Special Measures Order: Alert Levels and Restrictions in Newfoundland and Labrador

| **Alert Level** | **Restrictions** |
|---|---|
| Alert Level 1 (ALS-1) | The province will consider lifting long-term restrictions. |
| Alert Level 2 (ALS-2) | • Out-of-region travel for amateur sports, arts, and recreation, including competitions, tournaments, and training camps, is prohibited.<br>• Arenas, gyms, fitness facilities, and similar venues may operate at 50% capacity, including participants and spectators, provided physical distancing is maintained.<br>• Religious, spiritual, and cultural ceremonies, including funerals, burials, and weddings, are limited to 50% capacity if proof of vaccination is verified; otherwise, they are limited to 25% capacity.<br>• Bars and restaurants may operate at 50% capacity, while buffets are prohibited.<br>• Cinemas, performance spaces, bingo halls, and other indoor gatherings may operate at 50% capacity. Large venues (500+ capacity) may exceed this limit if an approved operating plan is in place.<br>• Private gatherings in homes or properties are limited to 25 people. |

| Alert Level | Restrictions |
|---|---|
| Alert Level 3 (ALS-3) | • Stay home as much as possible; household bubbles limited to 10 close contacts.<br>• Religious and cultural gatherings limited to 20 people; wakes prohibited.<br>• Gyms, pools, and fitness facilities can open, but team sports and performance spaces are closed.<br>• Retail stores can increase capacity; restaurants open at 50%, bars and lounges remain closed.<br>• Limited health care services and visitation allowed.<br>• Cinemas, bingo halls, and performance spaces remain closed.<br>Note: the modifications below were done when it was reintroduced.<br>• Closed businesses: Cinemas, performance spaces, and establishments primarily serving alcohol under the Liquor Control Act.<br>• Open with restrictions: Gyms, fitness facilities, dance studios, arenas, and similar venues may operate with a maximum of 100 people or 50% capacity.<br>• Group sports, recreation, and arts activities are suspended, except for team training and household-only lessons or performances.<br>• Restaurants may operate at 50% capacity for in-person. Buffets are prohibited, and bingo halls must close.<br>• Wakes are prohibited, with limited visitation allowed under physical distancing. Funerals, weddings, and ceremonies are limited to 100 people or 50% capacity with proof of vaccination, or 25% capacity without it.<br>• Retail stores, personal service establishments, and private health clinics may remain open at reduced capacity, following public health guidelines.<br>• Isolated or quarantined individuals must stay on their property/unit, avoid common spaces, and leave only for medical attention.<br>• Households may gather only with the same 20 close contacts, excluding work, school, and custody arrangements. |
| Alert Level 4 (ALS-4) | • Closed: Cinemas, performance spaces, and alcohol-serving establishments under the Liquor Control Act.<br>• Open with restrictions: Gyms, fitness facilities, dance studios, and arenas at max 50 people or 25% capacity, with household distancing.<br>• Amateur group sports, recreation, arts, and cultural activities are suspended, except for individual activities and household-only lessons or performances.<br>• Restaurants may operate at 50% capacity with 2-meter distancing and 6 people per table; buffets and formal event dancing (except ceremonial dances) are prohibited. Bingo halls remain closed.<br>• Wakes are prohibited, but limited visitation is allowed one household bubble at a time. Funerals, weddings, and ceremonies are limited to 50 people or 25% capacity, whichever is lower, with physical distancing.<br>• Indoor gatherings by businesses or organizations follow the same 50-person or 25% capacity limit, except large venues (500+) may exceed 25% capacity with an approved plan.<br>• Household gatherings are limited to the same 10 close and consistent contacts, except for work, school, and custody arrangements.<br>• Private health clinics, retail stores, and personal service establishments may remain open at reduced capacity, following public health guidelines and physical distancing.<br>• Individuals in self-isolation or quarantine must remain in their unit, avoid common spaces, and may only leave for medical attention. |

| Alert Level | Restrictions |
|---|---|
| Alert Level 5 (ALS-5) | • Gatherings, burials, and weddings limited to five people. Funerals are prohibited.<br>• Non-urgent health care is suspended, and private health clinics (except those of physicians or nurse practitioners) must close to non-urgent care.<br>• Non-essential retail businesses must close, except those providing life, health, or personal safety services for individuals and animals.<br>• Gyms, recreational facilities, performance spaces, cinemas, campsites, and playgrounds must close. |

##### S1.3 K–12 school operational status

In addition to the scheduled winter break, K–12 schools closed two days earlier on December 20, 2021, following recommendations from the Department of Health and Community Services (6). Schools remained closed for three additional weeks, with in-person learning resuming only on January 25, 2022 (7; 8). These intervals overlapped with the implementation of ALS-3 and ALS-4 restrictions as summarized in Table S2.

**Table S2**: **Time periods for alert levels and K–12 school closures**

| ALS Level | K–12 Status | Date Interval |
|---|---|---|
| ALS-2 | Open | Dec 15– Dec 19 (2021) |
| ALS-2 | Closed | Dec 20–24 (2021) |
| ALS-3 | Closed | Dec 25 – Jan 3 (2022) |
| ALS-4 | Closed | Jan 4 – Jan 25 (2022) |
| ALS-4 | Open | Jan 26 – Feb 7 (2022) |
| ALS-3 | Open | Feb 8 – Mar 14 (2022) |
| No-ALS | Open | Mar 15 – May 22 (2022) |

##### S1.4 PCR Testing Eligibility Criteria

**Table S3**: **COVID-19 PCR testing eligibility criteria in Newfoundland and Labrador during the Omicron wave.**

| Date | Testing Eligibility Criteria | Ref. | Label |
|---|---|---|---|
| 2021-12-15 | • Anyone with $\geq$1 COVID-19 symptom, regardless of vaccination status.<br>• Returning post-secondary students (domestic/international) tested upon arrival.<br>• Testing of contacts as part of case/contact tracing, including exposure notifications (e.g., flights, venues). | (9) | T1 |
| 2022-01-03 | • PCR recommended for close contacts without symptoms.<br>• PCR recommended for symptomatic individuals not identified as close contacts. | (10) | T2 |

| Date | Testing Eligibility Criteria | Ref. | Label |
|---|---|---|---|
| 2022-01-24 | • PCR no longer required for household contacts or unvaccinated non-household contacts.<br>• Still recommended for vaccinated non-household contacts without symptoms and symptomatic individuals not identified as close contacts. | (11) | T3 |
| 2022-02-25 | • Asymptomatic vaccinated household contacts: modified isolation (5 days) + PCR ≥72h post-exposure.<br>• Asymptomatic partially/unvaccinated household contacts: 7-day isolation + PCR ≥72h post-exposure.<br>• Symptomatic household contacts (all): PCR eligible.<br>• Symptomatic non-household contacts (all): immediate test (PCR or rapid). | (12) | T4 |
| 2022-03-17 | • PCR restricted to symptomatic individuals at high risk for severe COVID-19:<br>– ≥60 years, children <2 years, Indigenous adults (18+).<br>– Frontline healthcare workers, pregnant individuals.<br>– Those in congregate settings, immunocompromised persons. | (13) | T5 |

#### S1.5 Model equations and control reproduction number

The model in the main text is an SEAIR framework stratified into three vaccination cohorts describing SARS-CoV-2 transmission under an alert level system, periods of school closures, and given an ongoing vaccination program. The cohorts represent unvaccinated or single-dose individuals ($S_1$), two-dose ($V_2$), and three-or-more–dose ($V_3$) individuals. Within each cohort, individuals move through the same stages: susceptible → exposed ($E_j$) → infectious (asymptomatic, $A_j$, or symptomatic, $I_j$) → recovered ($R_j$). Infection risk is shared across cohorts via a common force of infection, with asymptomatic infectiousness reduced by $\zeta$ relative to symptomatic cases. Susceptibility is scaled by cohort-specific factors $\kappa_j$.

Vaccination was introduced into the model as an externally prescribed flow using observed vaccination data(14). The vaccination rate parameters $\nu_2(t)$ (to two doses) and $\nu_3(t)$ (to three or more doses) describe the rate of vaccination each day. These flows are modelled by the functions, $f_2(S_1)$ and $f_3(V_2)$, to ensure that vaccination does not exceed the number of eligible individuals in each cohort and to maintain numerical stability. See Section S1.7 for a graph of $f_2(S_1)$ and $f_3(V_2)$.

The complete set of continuous-time ordinary differential equations describing these flows, corresponding to the flowchart in the main text, is given below.

$$\lambda_d(t) = \beta(t)\,\frac{\zeta\,(A_1+A_2+A_3) + (I_1+I_2+I_3)}{N}.$$

$$f_2(S_1) = \frac{\nu_2(t)S_1}{\nu_2(t) + S_1}, \qquad f_3(V_2) = \frac{\nu_3(t)V_2}{\nu_3(t) + V_2}.$$

**Cohort 1 (unvaccinated)**
$$\begin{cases} \dfrac{dS_1}{dt} = -\kappa_1 \lambda_d S_1 - f_2(S_1), \\ \dfrac{dE_1}{dt} = \kappa_1 \lambda_d S_1 - \sigma E_1, \\ \dfrac{dI_1}{dt} = \mu\, \sigma E_1 - \gamma_i I_1, \\ \dfrac{dA_1}{dt} = (1-\mu)\, \sigma E_1 - \gamma_a A_1, \\ \dfrac{dR_1}{dt} = \gamma_i I_1 + \gamma_a A_1. \end{cases}$$

**Cohort 2 (two doses)**
$$\begin{cases} \dfrac{dV_2}{dt} = f_2(S_1) - \kappa_2 \lambda_d V_2 - f_3(V_2), \\ \dfrac{dE_2}{dt} = \kappa_2 \lambda_d V_2 - \sigma E_2, \\ \dfrac{dI_2}{dt} = \mu\, \sigma E_2 - \gamma_i I_2, \\ \dfrac{dA_2}{dt} = (1-\mu)\, \sigma E_2 - \gamma_a A_2, \\ \dfrac{dR_2}{dt} = \gamma_i I_2 + \gamma_a A_2. \end{cases}$$

**Cohort 3 (3+ doses)**
$$\begin{cases} \dfrac{dV_3}{dt} = f_3(V_2) - \kappa_3 \lambda_d V_3, \\ \dfrac{dE_3}{dt} = \kappa_3 \lambda_d V_3 - \sigma E_3, \\ \dfrac{dI_3}{dt} = \mu\, \sigma E_3 - \gamma_i I_3, \\ \dfrac{dA_3}{dt} = (1-\mu)\, \sigma E_3 - \gamma_a A_3, \\ \dfrac{dR_3}{dt} = \gamma_i I_3 + \gamma_a A_3. \end{cases}$$

We use a fixed population size, $N = 510,550$, based on Statistics Canada's 2021 Census population for NL. We add the number of individuals in the respective compartments across the three cohorts to obtain the total number of individuals per disease state in the population at any given time, $t$, i.e., $E = E_1 + E_2 + E_3$. The community transmission dynamics of Omicron in the whole province is summarized as:

$$\begin{cases} \dfrac{dS}{dt} = \dfrac{dS_1}{dt} + \dfrac{dV_2}{dt} + \dfrac{dV_3}{dt}, \\ \dfrac{dE}{dt} = \dfrac{dE_1}{dt} + \dfrac{dE_2}{dt} + \dfrac{dE_3}{dt}, \\ \dfrac{dA}{dt} = \dfrac{dA_1}{dt} + \dfrac{dA_2}{dt} + \dfrac{dA_3}{dt}, \\ \dfrac{dI}{dt} = \dfrac{dI_1}{dt} + \dfrac{dI_2}{dt} + \dfrac{dI_3}{dt}, \\ \dfrac{dR}{dt} = \dfrac{dR_1}{dt} + \dfrac{dR_2}{dt} + \dfrac{dR_3}{dt}. \end{cases} \tag{1}$$

#### The control reproduction number, $R_c(t)$

To derive the control reproduction number, $R_c(t)$, we apply the next-generation matrix method (15). We define the infected-state vector as

$$x = \left(E_1, A_1, I_1,\ E_2, A_2, I_2,\ E_3, A_3, I_3\right)^\top.$$

corresponding to exposed, asymptomatic, and symptomatic individuals in each vaccination cohort.

At the disease-free equilibrium (DFE), all infected compartments are zero, and the susceptible pools are $S_j \in \{ S_1, V_2, V_3\}$ with population proportions

$$p_j = \frac{S_j}{N}.$$

New infections occur only in the exposed compartments. For cohort $j$, the rate of occurrence of new infections is given as

$$\mathcal{F}_{E_j} = \kappa_j \lambda_d(t)\, S_j,$$

depending on vaccination status. Linearizing the force of infection around the DFE gives

$$\lambda_d(t) = \beta(t) \frac{\zeta(A_1{+}A_2{+}A_3) + (I_1{+}I_2{+}I_3)}{N}.$$

The Jacobian $F = [\partial \mathcal{F}_i/\partial x_k]_{\mathrm{DFE}}$ has nonzero entries only in rows corresponding to $E_1, E_2, E_3$ and columns corresponding to $A_1, A_2, A_3, I_1, I_2, I_3$. For any $j, k \in \{1, 2, 3\}$,

$$\frac{\partial \mathcal{F}_{E_j}}{\partial A_k}\Big|_{\mathrm{DFE}} = \kappa_j\, \beta(t)\, \frac{\zeta\, S_j}{N}, \qquad \frac{\partial \mathcal{F}_{E_j}}{\partial I_k}\Big|_{\mathrm{DFE}} = \kappa_j\, \beta(t)\, \frac{S_j}{N}.$$

Using the ordering $(E_1, A_1, I_1, E_2, A_2, I_2, E_3, A_3, I_3)$, the full $9 \times 9$ matrix is

$$F(t) = \begin{pmatrix}
0 & \frac{\kappa_1\beta(t)\zeta S_1}{N} & \frac{\kappa_1\beta(t)S_1}{N} & 0 & \frac{\kappa_1\beta(t)\zeta S_1}{N} & \frac{\kappa_1\beta(t)S_1}{N} & 0 & \frac{\kappa_1\beta(t)\zeta S_1}{N} & \frac{\kappa_1\beta(t)S_1}{N} \\
0 & 0 & 0 & 0 & 0 & 0 & 0 & 0 & 0 \\
0 & 0 & 0 & 0 & 0 & 0 & 0 & 0 & 0 \\
0 & \frac{\kappa_2\beta(t)\zeta V_2}{N} & \frac{\kappa_2\beta(t)V_2}{N} & 0 & \frac{\kappa_2\beta(t)\zeta V_2}{N} & \frac{\kappa_2\beta(t)V_2}{N} & 0 & \frac{\kappa_2\beta(t)\zeta V_2}{N} & \frac{\kappa_2\beta(t)V_2}{N} \\
0 & 0 & 0 & 0 & 0 & 0 & 0 & 0 & 0 \\
0 & 0 & 0 & 0 & 0 & 0 & 0 & 0 & 0 \\
0 & \frac{\kappa_3\beta(t)\zeta V_3}{N} & \frac{\kappa_3\beta(t)V_3}{N} & 0 & \frac{\kappa_3\beta(t)\zeta V_3}{N} & \frac{\kappa_3\beta(t)V_3}{N} & 0 & \frac{\kappa_3\beta(t)\zeta V_3}{N} & \frac{\kappa_3\beta(t)V_3}{N} \\
0 & 0 & 0 & 0 & 0 & 0 & 0 & 0 & 0 \\
0 & 0 & 0 & 0 & 0 & 0 & 0 & 0 & 0
\end{pmatrix}$$

All remaining transitions among infected compartments are collected in the matrix $V$, which captures progression and recovery. Because cohorts do not exchange infection states directly, $V$ is block diagonal, with identical lower-triangular blocks corresponding to each cohort. For a single cohort $j$, restricted to $(E_j, A_j, I_j)$,

$$\begin{aligned}
\dot{E}_j &= \mathcal{F}_{E_j} - \sigma E_j, \\
\dot{A}_j &= 0 - \big(\gamma_a A_j - (1-\mu)\sigma E_j\big), \\
\dot{I}_j &= 0 - \big(\gamma_i I_j - \mu\sigma E_j\big).
\end{aligned}$$

The corresponding Jacobian block is

$$V_{\mathrm{blk}} = \begin{pmatrix} \sigma & 0 & 0 \\ -(1-\mu)\sigma & \gamma_a & 0 \\ -\mu\sigma & 0 & \gamma_i \end{pmatrix}$$

resulting in the full matrix, which is block diagonal,

$$V = \mathrm{diag}(V_{\mathrm{blk}}, V_{\mathrm{blk}}, V_{\mathrm{blk}}).$$

Since $V_{\text{blk}}$ is lower triangular, its inverse is

$$V^{-1} = \begin{pmatrix} \frac{1}{\sigma} & 0 & 0 & 0 & 0 & 0 & 0 & 0 & 0 \\ \frac{1-\mu}{\gamma_a} & \frac{1}{\gamma_a} & 0 & 0 & 0 & 0 & 0 & 0 & 0 \\ \frac{\mu}{\gamma_i} & 0 & \frac{1}{\gamma_i} & 0 & 0 & 0 & 0 & 0 & 0 \\ 0 & 0 & 0 & \frac{1}{\sigma} & 0 & 0 & 0 & 0 & 0 \\ 0 & 0 & 0 & \frac{1-\mu}{\gamma_a} & \frac{1}{\gamma_a} & 0 & 0 & 0 & 0 \\ 0 & 0 & 0 & \frac{\mu}{\gamma_i} & 0 & \frac{1}{\gamma_i} & 0 & 0 & 0 \\ 0 & 0 & 0 & 0 & 0 & 0 & \frac{1}{\sigma} & 0 & 0 \\ 0 & 0 & 0 & 0 & 0 & 0 & \frac{1-\mu}{\gamma_a} & \frac{1}{\gamma_a} & 0 \\ 0 & 0 & 0 & 0 & 0 & 0 & \frac{\mu}{\gamma_i} & 0 & \frac{1}{\gamma_i} \end{pmatrix}$$

The next-generation matrix is defined as

$$K(t) = F(t)\,V^{-1},$$

where $F(t)$ encodes the rate at which new infections are generated in each cohort and $V^{-1}$ captures the expected time spent in infectious states before removal. Due to the shared force of infection $\lambda_d(t)$ and additive contributions from all infectious compartments, $K(t)$ has rank one. The product of $F(t)$ by $V^{-1}$ collapses the exposed, asymptomatic, and symptomatic pathways into a single per-infection contribution proportional to the mean infectious period,

$$\frac{\mu}{\gamma_i} + \frac{(1-\mu)\zeta}{\gamma_a},$$

while weighting each vaccination cohort by its relative susceptibility $\kappa_j$ and its population proportion $p_j$. The dominant eigenvalue (spectral radius) of $K(t)$ therefore simplifies to

$$\mathcal{R}_c(t) = \beta(t)\left(\sum_{j=1}^{3} p_j\kappa_j\right)\left(\frac{\mu}{\gamma_i} + \frac{(1-\mu)\zeta}{\gamma_a}\right).$$

Finally, averaging the time-varying transmission rate over the interval $[t_a, t_b]$:

$$\bar{\beta}[t_a, t_b] = \frac{1}{t_b - t_a}\int_{t_a}^{t_b} \beta(t)\,dt,$$

yields the control reproduction number:

$$\mathcal{R}_c(t) = \bar{\beta}[t_a, t_b]\sum_{j=1}^{3} p_j\kappa_j\left(\frac{\mu}{\gamma_i} + \frac{(1-\mu)\zeta}{\gamma_a}\right).$$

#### S1.6 Priors for Bayesian estimation of fitted parameters

Model parameters and their values are provided in Table 1 of the main text. We estimated some parameters by fitting them with weakly informative priors. These priors were approximately centered at $m$ based on literature values, with standard deviation, $s$, and allowed the 95% prior intervals to represent plausible ranges. For each rate parameter, $\theta_{\text{rate}} \in \{\gamma_a, \gamma_i, \sigma\}$, this range is [1/11, 1/8], [1/8, 1/6], and [1/4, 1/2] days$^{-1}$ respectively. For the proportion parameters $\theta_{\text{prop}} \in \{\kappa_2, \kappa_3\}$, we define their plausible ranges as [0.85, 0.96] and [0.20, 0.40], respectively. We used Gaussian priors on the log scale for the $\theta_{\text{rate}}$ parameters to ensure positivity. We used Gaussian priors on the logit scale for the $\theta_{\text{prop}}$ parameters to ensure that these unitless parameters lie between zero and one. Throughout, we use the notation $\mathcal{N}(m, s^2)$ to denote the Normal distribution with mean $m$ and variance $s^2$. Here, "lower" and "upper" denote the bounds of the defined parameter value ranges. Specifically, for rate parameters we used

$$\log(\theta_{\text{rate}}) \sim \mathcal{N}(m,\, s^2), \tag{2}$$

with

$$m = \frac{\log(\text{lower}) + \log(\text{upper})}{2}, \qquad s = \frac{\log(\text{upper}) - \log(\text{lower})}{2 \times 1.96}.$$

For proportions we used

$$\text{logit}(\theta_{\text{prop}}) \sim \mathcal{N}(m,\, s^2), \tag{3}$$

with

$$m = \frac{\text{logit}(\text{lower}) + \text{logit}(\text{upper})}{2}, \qquad s = \frac{\text{logit}(\text{upper}) - \text{logit}(\text{lower})}{2 \times 1.96}.$$

Fig S2 summarizes the posterior summaries of the fitted parameters. Posterior point estimates and 95% uncertainty intervals indicate that all parameters are well informed by the data, with uncertainty substantially reduced relative to the corresponding prior ranges. The rate parameters as well as the proportion parameters are all identifiable and remain within epidemiologically plausible bounds.

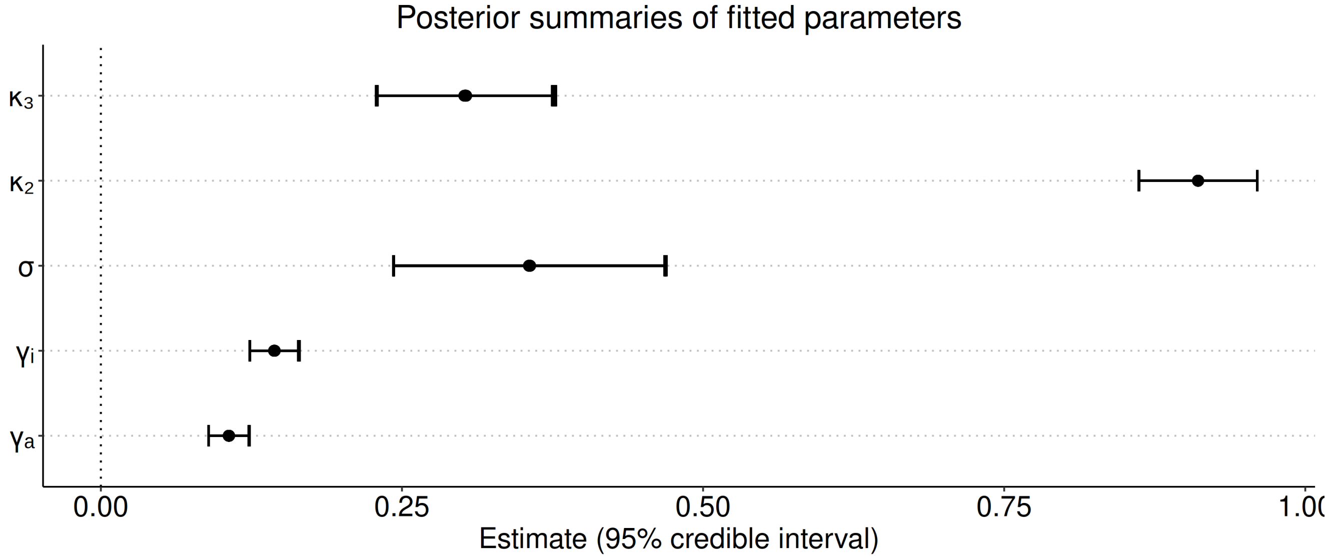


**Fig. S2**: **Posterior summaries of fitted model parameters.** Points denote posterior point estimates and horizontal bars indicate 95% credible intervals. Parameters shown are the relative susceptibility of double-vaccinated individuals ($\kappa_2$), the relative susceptibility of boosted individuals ($\kappa_3$), the rate of progression from exposed to infectious ($\sigma$), the recovery rate of symptomatic infections ($\gamma_i$), and the recovery rate of asymptomatic infections ($\gamma_a$).

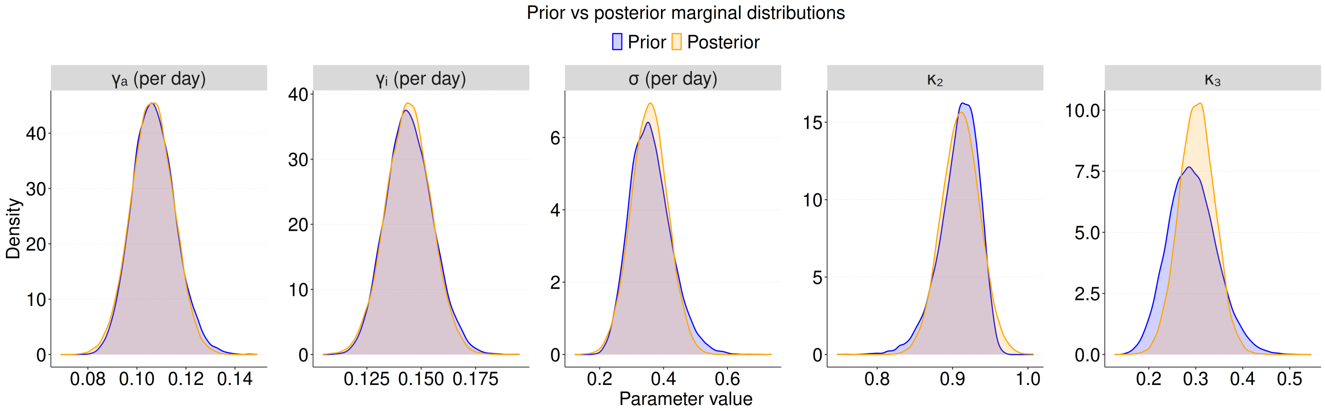


**Fig. S3**: **Prior and posterior marginal distributions of fitted model parameters.** Blue curves show prior distributions and orange curves show posterior distributions. Panels correspond to the recovery rate of asymptomatic infections ($\gamma_a$, per day), the recovery rate of symptomatic infections ($\gamma_i$, per day), the rate of progression from exposed to infectious ($\sigma$, per day), the relative susceptibility of double-vaccinated individuals ($\kappa_2$), and the relative susceptibility of boosted individuals ($\kappa_3$).

#### S1.7 Vaccination Data

Publicly available COVID-19 vaccination data for Newfoundland and Labrador were obtained from the Public Health Agency of Canada (PHAC) website (14). These data are reported as cumulative vaccination counts at a weekly resolution. To incorporate vaccination into the daily-time-step transmission model, the weekly cumulative series for second doses and booster doses were first converted to smooth daily cumulative trajectories using spline-based interpolation. Daily vaccination counts were then obtained by differencing the interpolated cumulative series.

As shown in Fig. S4, the resulting daily numbers of individuals receiving second doses (left panel) and booster doses (right panel) were used directly as time-varying inputs to the model. These daily vaccination counts were implemented as vaccination rates, ensuring that the number of individuals transitioning into the double-vaccinated and boosted compartments on each day exactly matched the empirical vaccination rollout. This approach allowed the model to accurately reflect how the immunity profile of the population changed over time.

We note that NL began its COVID-19 booster dose campaign on November 8, 2021, initially restricting eligibility to individuals who had received their second dose at least six months earlier. On December 22, 2021 this eligibility was changed to individuals who had received their second dose at least five months (22 weeks) prior. The booster dose campaign occurred rapidly and there are gaps in the initial reporting. According to the House of Assembly report, 'by January 11, 2022, at least 25 percent of those aged 12 and older had received a third dose', although this figure was considered a conservative estimate because of 'the increase in paper-based clinics and required back entry of those records'. Such operational constraints, particularly delays in digitizing manually collected records, likely contributed to discrepancies in the timing of booster coverage estimates.

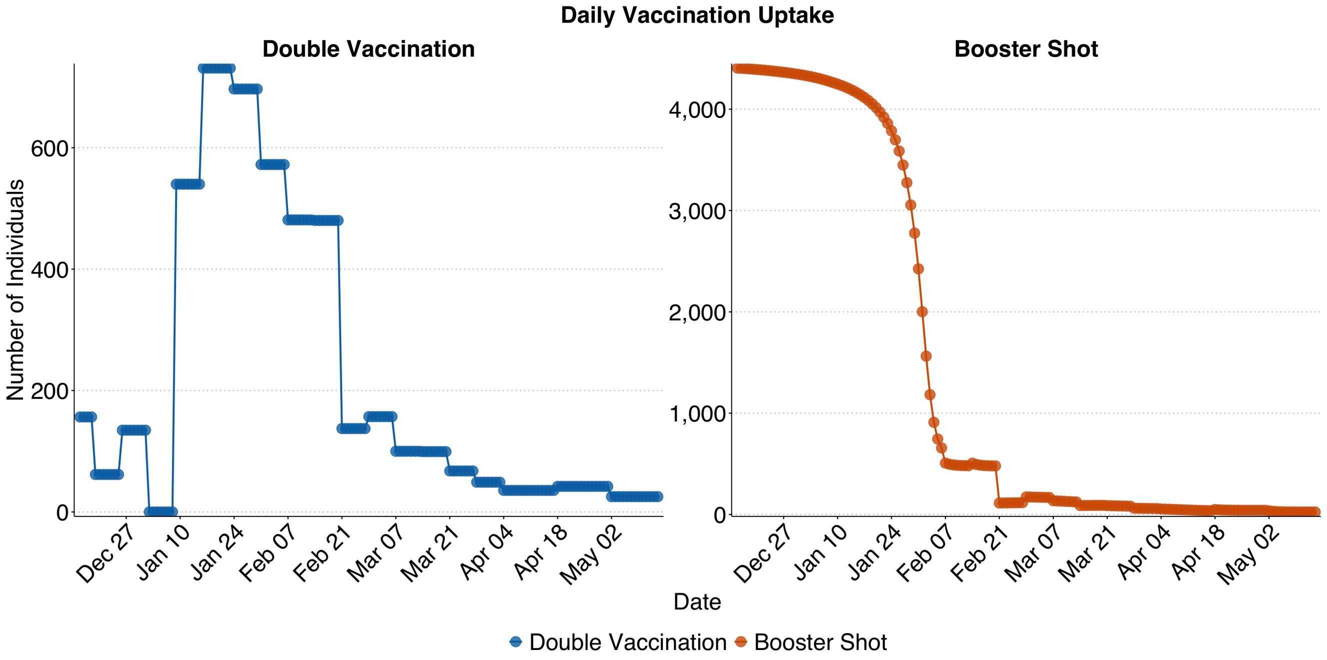


**Fig. S4**: **Daily COVID-19 vaccination uptake in Newfoundland and Labrador during the Omicron wave.** Points show daily vaccination counts derived from PHAC-reported weekly cumulative data for second doses (left panel) and booster doses (right panel) between December 15, 2021, and May 22, 2022. $f_2(S_1)$ and $f_3(V_3)$ are shown as the lines representing the daily vaccination rates, which were implemented as time-varying inputs in the model.

#### S1.8 Sensitivity analysis

We evaluated the sensitivity of model outputs to uncertainty in two fixed parameters: the symptomatic proportion, $\mu$, and the relative infectivity of individuals with asymptomatic infections, $\zeta$. These parameters were selected because the other fixed parameters in the model are either reference values ($\kappa_1$), estimated with a higher level of certainty ($N$), or estimated directly from data ($v_1(t), v_2(t)$). Fitted parameters were not considered for the sensitivity analysis because the uncertainty in these parameters is described by their posterior distributions.

For the sensitivity analysis, we used a full factorial design and considered $\mu$ for a range of values between 0.45 and 0.85, consistent with systematic reviews estimating that approximately 17–45% of SARS-CoV-2 infections remain asymptomatic, corresponding to a symptomatic proportion of 55–85% (16; 17). We considered a range of $\zeta$ values between 0.40 and 1.00, reflecting empirical evidence that asymptomatic individuals contribute substantially to transmission, with infectiousness ranging from moderately reduced to comparable with that of symptomatic cases (18–20). We considered all 25 combinations of 5 equally spaced values of $\mu$ and $\zeta$ spanning these stated ranges.

For each combination of $\mu$ and $\zeta$, the model was fully recalibrated using the same data sources, likelihood structure, and estimation procedure as in the primary analysis. For each of the 25 values of $(\mu, \zeta)$ that were considered for the sensitivity analysis, we fit values of $\gamma_a$, $\gamma_i$, $\sigma$, $\kappa_2$, and $\kappa_3$. For each of the 25 values considered, the estimates of the fitted parameter values was consistent and within biologically plausible ranges (Fig S5). No evidence of numerical instability or boundary convergence was observed, indicating that model calibration was well identified across the explored parameter space.

The main conclusions of our manuscript concern our estimates of the time-varying control reproduction number, $\mathcal{R}_c(t)$. To understand the sensitivity of our conclusions to uncertainty in $\mu$ and $\zeta$ we estimated the control reproduction number for each value $(\mu, \zeta)$ pair in the sensitivity analysis (Fig S6). The control reproduction number varied $R_c(t)$ minimally for the different values of $(\mu, \zeta)$ that we considered. The estimated $\mathcal{R}_c(t)$ dynamics have nearly identical values at all points in time for all the values of $(\mu, \zeta)$ considered, such that Figs 5 and 6 in the main text would be nearly identical had we chosen different values of $\mu$ and $\zeta$ in Table 1 of the main text. Fig 5 in the main text is analogous to Fig S5 but for the parameter values in Table 1 and with 95% confidence intervals. Fig 6 in the main text aggregates the $R_c(t)$ values for alert levels and K–12 school status as open or closed. Overall, the main

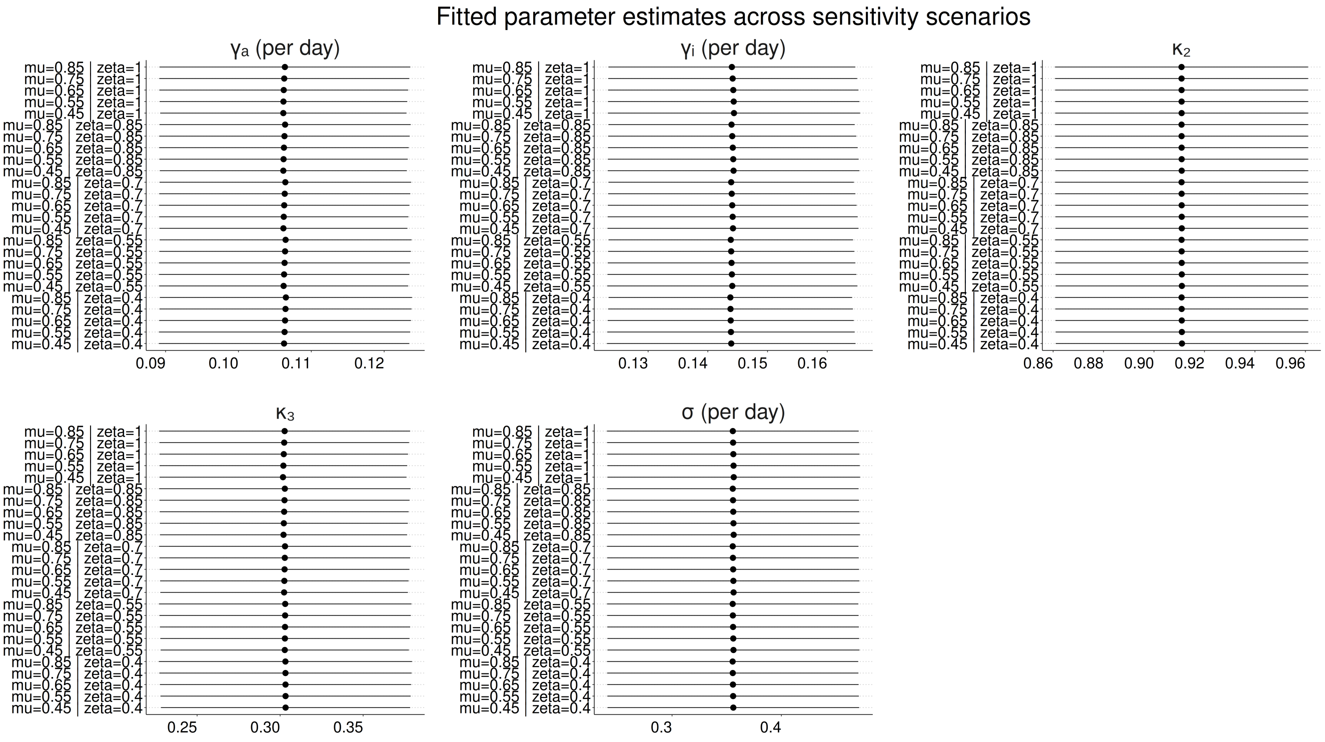


**Fig. S5**: **Sensitivity of fitted epidemiological parameter values to different values of the symptomatic proportion, $\mu$, and asymptomatic infectivity, $\zeta$.** Posterior point estimates of fitted parameters ($\gamma_a$, $\gamma_i$, $\sigma$, $\kappa_2$, and $\kappa_3$) are shown for all 25 values of $\mu$ and $\zeta$ considered for the sensitivity analysis). Each point represents the fitted estimate for a $(\mu, \zeta)$ combination, with horizontal lines indicating the uncertainty in the fitted estimate. Parameter estimates remain consistent across the explored parameter space, indicating that model calibration is robust to uncertainty in $\mu$ and $\zeta$.

conclusions are robust to substantial uncertainty in the symptomatic proportion and the relative infectivity of asymptomatic infections.

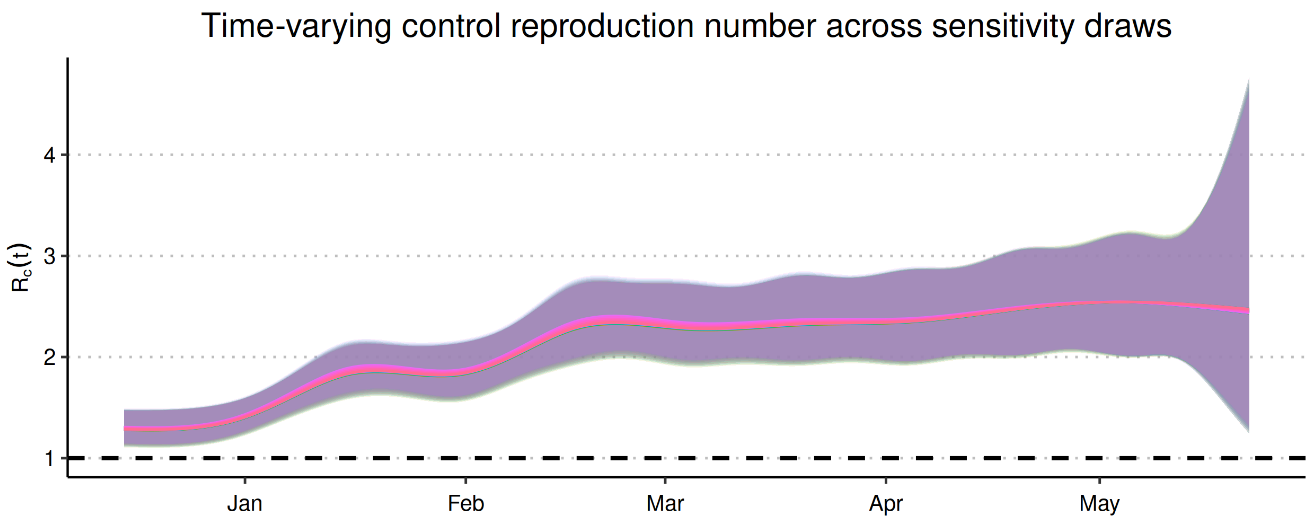


**Fig. S6**: **Time-varying control reproduction number across sensitivity draws.** The control reproduction number, $\mathcal{R}_c(t)$, was computed over time for the sensitivity analysis in which the symptomatic proportion ($\mu$) and the relative infectivity of asymptomatic infections ($\zeta$) were varied. The shaded band summarizes the range of $\mathcal{R}_c(t)$ values across all parameter combinations, and the solid curve shows the corresponding central tendency over time. The dashed horizontal line marks the epidemic threshold ($\mathcal{R}_c(t) = 1$).

#### S1.9 Restrictive PCR testing eligibility in Canadian provinces.

PCR was the primary diagnostic method used in Canada to confirm SARS-CoV-2 infection. The provincial and territorial governments determined the testing strategies and eligibility based on their epidemiological context and laboratory capacity. As the PCR testing capacity was exceeded shortly after the establishment of the Omicron variant, many provinces and territories changed the PCR test eligibility criteria. The dates in which different provinces implemented restrictive eligibility for PCR tests are summarized in Table S4, where restrictive eligibility is defined as requiring individuals to be symptomatic and high-risk or in frequent contact with high-risk groups.

**Table S4**: **Restrictive PCR testing eligibility in Canadian provinces.** Restrictive eligibility is defined as requiring individuals to be both symptomatic and either high-risk or in frequent contact with high-risk groups.

| **Province** | **Date introduced** | **Reference** | **Comments** |
|---|---|---|---|
| Alberta (AB) | Jun 30, 2022 | (21–23) | Close contacts redirected to at-home RAT; PCR testing only via referral and restricted to high-risk individuals (e.g., antiviral eligibility, pregnant symptomatic, First Nations/remote high-risk settings). |
| British Columbia (BC) | Sep 28, 2022 | (24; 25) | PCR testing limited to cases where results affect treatment; focused on immunocompromised, seniors without boosters, high-risk conditions, pregnant women, hospitalized, and antiviral-eligible individuals. |
| Manitoba (MB) | Apr 15, 2022 | (26; 27) | Provincial sites closed; PCR reserved for severe cases or high-risk settings (pre-op, immunocompromised, congregate, recent travellers, clinical decision-making). |

Continued on next page

**Table S4 – continued from previous page**

| **Province** | **Date introduced** | **Reference** | **Comments** |
|---|---|---|---|
| New Brunswick (NB) | Jan 4, 2022 | (28) | PCR testing by self-referral restricted to symptomatic high-risk groups (50+ or <2 years, healthcare and congregate-care workers or residents, immunocompromised individuals, pregnant people, the precarious shelter, and those requiring PCR tests for international travel) |
| Newfoundland & Labrador (NL) | Mar 17, 2022 | (29; 30) | PCR restricted to symptomatic high-risk groups (immunocompromised, pregnant, First Nations/Inuit/Métis adults, <2 years, seniors, congregate settings, HCWs). Earlier Jan 2022 changes excluded confirmatory testing for many close contacts. |
| Nova Scotia (NS) | Dec 24, 2021 | (31) | Most residents became ineligible for provincial PCR; eligibility restricted to high-risk groups and frequent contacts of high-risk individuals (e.g., patient-facing HCWs). |
| Prince Edward Island (PEI) | Not introduced | — | Appointment-based testing introduced Jun 8, 2022, but province did **not** adopt restrictive PCR eligibility. |
| Quebec (QC) | Jan 4, 2022 | (32–34) | PCR testing restricted to high-risk individuals and settings (symptomatic hospital patients, residents of congregate care, people transferring between medical facilities, detention centres, homeless shelters, and northern or remote communities). |
| Ontario (ON) | Apr 11, 2022 | (35; 36) | PCR prioritized to symptomatic high-risk groups (immunocompromised, ≥60 with <3 doses, comorbidities, pregnant, HCWs, first responders, congregate settings). Earlier on Dec 30, 2021, symptomatic residents were assumed positive and asymptomatic contacts were largely excluded. |
| Saskatchewan (SK) | Feb 7, 2022 | (37) | PCR restricted to priority at-risk groups (e.g., late-term pregnant, immunocompromised, LTC/personal care residents, Indigenous/remote communities, specific infant/parent cases). |

#### S1.10 Estimating underreporting ratio for Canadian provinces

We found that in early January 2022, reported case counts briefly exceeded seroprevalence in NL. Below we show that this has been a characteristic of Canadian seroprevalence data, particularly during periods of, or for provinces with, low infection prevalence. The reported COVID-19 case counts were obtained from the Public Health Agency of Canada (38), and provincial population sizes were taken from the 2021 Census conducted by Statistics Canada (39). Data were summarized for four intervals: July–December 2020, January–June 2021, July–December 2021, and January–June 2022. Estimates of infection-induced seroprevalence were provided by the COVID-19 Immunity Task Force (CITF) (40), representing the proportion of the population with evidence of prior infection. The underreporting ratio for each interval was then calculated as the percentage change in seroprevalence divided by the percentage change in reported infections per capita.

**Table S5**: **Estimated under-reporting of COVID-19 cases by province and half-year period.** Reported case counts are cumulative totals and underreporting ratios are calculated as the percentage change in N-antibody seropositivity divided by the percentage change in cumulative cases per capita. Estimates of the under-reporting ratio less than one are bolded and indicate more reported cases than infections estimated by serosurveillance.

| Province (population) | Period | Cum cases (start) | Cum cases (end) | Δ cum cases | Δ% cases per capita pct | serop (start) | serop (end) | Δ% serop | Under-reporting ratio |
|---|---|---|---|---|---|---|---|---|---|
| Alberta (4,262,635) | 2020: Jul–Dec | 8,286 | 97,407 | 89,121 | 2.1 | 0.8% (0.4–1.4) | 2.6% (1.9–3.3) | 1.8 | **0.9** |
| | 2021: Jan–Jun | 104,633 | 231,810 | 127,177 | 3.0 | 2.7% (2.0–3.5) | 6.2% (5.0–7.6) | 3.5 | 1.2 |
| | 2021: Jul–Dec | 232,163 | 352,873 | 120,710 | 2.8 | 6.3% (5.1–7.7) | 11.3% (9.5–13.1) | 5.0 | 1.8 |
| | 2022: Jan–Jun | 373,555 | 588,311 | 214,756 | 5.0 | 12.7% (11.0–14.7) | 55.2% (55.2–58.2) | 42.5 | 8.5 |
| British Columbia (5,000,879) | 2020: Jul–Dec | 2,947 | 49,568 | 46,621 | 1.0 | 0.6% (0.3–1.1) | 1.8% (1.3–2.4) | 1.2 | 1.2 |
| | 2021: Jan–Jun | 53,162 | 147,475 | 94,313 | 2.9 | 1.9% (1.4–2.5) | 5.2% (4.2–6.2) | 3.3 | 1.1 |
| | 2021: Jul–Dec | 147,735 | 238,210 | 90,475 | 4.8 | 5.3% (4.3–6.4) | 9.1% (7.6–10.7) | 3.8 | **0.8** |
| | 2022: Jan–Jun | 258,882 | 374,594 | 115,712 | 7.5 | 10.5% (9.0–12.2) | 43.7% (41.3–46.3) | 33.2 | 4.4 |
| Manitoba (1,342,153) | 2020: Jul–Dec | 325 | 23,624 | 23,299 | 1.8 | 0.8% (0.4–1.4) | 3.5% (2.6–4.7) | 2.7 | 1.5 |
| | 2021: Jan–Jun | 25,026 | 55,878 | 30,852 | 4.2 | 3.6% (2.7–4.8) | 5.5% (4.5–6.7) | 1.9 | **0.5** |
| | 2021: Jul–Dec | 56,353 | 74,358 | 18,005 | 5.5 | 5.6% (4.6–6.8) | 10.1% (8.5–11.9) | 4.5 | **0.8** |
| | 2022: Jan–Jun | 80,096 | 145,498 | 65,402 | 10.8 | 11.5% (9.8–13.4) | 50.5% (47.5–53.5) | 39.0 | 3.6 |
| New Brunswick (775,610) | 2020: Jul–Dec | 165 | 590 | 425 | 0.1 | 0.5% (0.2–0.8) | 1.1% (0.7–1.6) | 0.6 | 1.2 |
| | 2021: Jan–Jun | 611 | 2,324 | 1,713 | 0.3 | 1.1% (0.8–1.6) | 1.8% (1.3–2.4) | 0.7 | 2.3 |
| | 2021: Jul–Dec | 2,335 | 11,896 | 9,561 | 1.5 | 1.8% (1.3–2.4) | 3.1% (2.4–4.0) | 1.3 | **0.9** |
| | 2022: Jan–Jun | 15,069 | 67,725 | 52,656 | 8.7 | 3.5% (2.7–4.4) | 44.5% (41.1–47.8) | 41.0 | 4.7 |
| Newfoundland & Labrador (510,550) | 2020: Jul–Dec | 261 | 384 | 123 | 0.1 | 0.5% (0.26–0.9) | 1.1% (0.8–1.6) | 0.6 | 6 |
| | 2021: Jan–Jun | 390 | 1,384 | 994 | 0.3 | 1.2% (0.8–1.7) | 2.3% (1.7–3.0) | 1.1 | **3.7** |
| | 2021: Jul–Dec | 1,387 | 2,512 | 1,125 | 0.5 | 2.3% (1.7–3.1) | 2.9% (2.2–3.9) | 0.6 | **1.2** |
| | 2022: Jan–Jun | 4,597 | 47,773 | 43,176 | 9.4 | 3.2% (2.4–4.2) | 45.5% (41.7–49.5) | 42.3 | 4.5 |
| Nova Scotia (969,383) | 2020: Jul–Dec | 1,064 | 1,469 | 405 | 0.2 | 0.4% (0.2–0.8) | 1.1% (0.7–1.5) | 0.7 | 3.5 |

*Continued on next page*

| **Province (population)** | **Period** | **Cum cases (start)** | **Cum cases (end)** | **Δ cum cases** | **Δ% cases per capita pct** | **serop (start)** | **serop (end)** | **Δ% serop** | **Under-reporting ratio** |
|---|---|---|---|---|---|---|---|---|---|
| | 2021: Jan–Jun | 1,497 | 5,820 | 4,323 | 0.6 | 1.1% (0.7–1.6) | 2.0% (1.5–2.6) | 0.9 | 1.5 |
| | 2021: Jul–Dec | 5,847 | 14,301 | 8,454 | 1.5 | 2.0% (1.5–2.6) | 3.3% (2.5–4.2) | 1.3 | **0.9** |
| | 2022: Jan–Jun | 19,175 | 106,633 | 87,458 | 11.0 | 3.8% (3.0–4.8) | 40% (36.9–43.2) | 36.2 | 3.3 |
| Ontario (14,223,942) | 2020: Jul–Dec | 35,897 | 173,501 | 137,604 | 1.2 | 0.8% (0.4–1.3) | 2.3% (1.7–3.0) | 1.5 | 1.3 |
| | 2021: Jan–Jun | 193,960 | 544,796 | 350,836 | 3.8 | 2.4% (1.8–3.2) | 4.8% (3.8–5.9) | 2.4 | **0.6** |
| | 2021: Jul–Dec | 546,268 | 705,514 | 159,246 | 5.0 | 4.9% (3.9–6.0) | 7.9% (6.4–9.5) | 3.0 | **0.6** |
| | 2022: Jan–Jun | 807,841 | 1,329,416 | 521,575 | 9.3 | 8.7% (7.4–10.6) | 47.3% (44.3–50.1) | 38.6 | 4.2 |
| Prince Edward Island (154,331) | 2020: Jul–Dec | 30 | 94 | 64 | 0.1 | 0.4% (0.2–0.7) | 1.0% (0.7–1.5) | 0.6 | 6.0 |
| | 2021: Jan–Jun | 97 | 207 | 110 | 0.1 | 1.1% (0.7–1.6) | 1.9% (1.3–2.6) | 0.8 | 8.0 |
| | 2021: Jul–Dec | 208 | 737 | 529 | 0.5 | 1.9% (1.4–2.6) | 2.9% (2.1–3.9) | 2.0 | 4.0 |
| | 2022: Jan–Jun | 1,447 | 40,460 | 39,013 | 26.2 | 3.2% (2.4–4.3) | 44% (40–48.1) | 40.8 | 1.6 |
| Quebec (8,501,833) | 2020: Jul–Dec | 48,735 | 186,323 | 137,588 | 2.2 | 0.8% (0.4–1.5) | 2.5% (1.8–3.5) | 1.7 | **0.8** |
| | 2021: Jan–Jun | 204,101 | 369,727 | 165,626 | 4.3 | 2.6% (1.8–3.7) | 5.5% (4.1–7.1) | 2.9 | **0.7** |
| | 2021: Jul–Dec | 370,242 | 547,902 | 177,660 | 6.4 | 5.6% (4.2–7.2) | 11.6% (8.5–15.3) | 6.0 | **0.9** |
| | 2022: Jan–Jun | 656,573 | 1,089,710 | 433,137 | 12.8 | 12.6% (9.5–16.2) | 49% (44.6–53.4) | 36.4 | 2.9 |
| Saskatchewan (1,132,505) | 2020: Jul–Dec | 796 | 14,615 | 13,819 | 1.3 | 0.5% (0.3–0.8) | 1.9% (1.3–2.6) | 1.4 | 1.1 |
| | 2021: Jan–Jun | 15,844 | 48,706 | 32,862 | 4.3 | 2.0% (1.4–2.8) | 4.5% (3.4–5.8) | 2.5 | **0.6** |
| | 2021: Jul–Dec | 48,931 | 83,086 | 34,155 | 7.3 | 4.6% (3.5–5.8) | 8.3% (6.8–9.9) | 3.7 | **0.5** |
| | 2022: Jan–Jun | 85,188 | 139,661 | 54,473 | 12.3 | 9.4% (7.9–11.1) | 50.8% (47.6–53.8) | 41.4 | 3.4 |
